# Understanding influences of care-seeking behaviors for diarrheal illnesses: A qualitative meta-synthesis

**DOI:** 10.1101/2025.07.08.25331122

**Authors:** Marissa H. Miller, Skye M. Hilbert, Erica N. Rosser, Laura Sinko, Elizabeth C. Lee, Kirsten E. Wiens

**Affiliations:** Department of Epidemiology and Biostatistics, Temple University College of Public Health, Philadelphia, USA; Department of International Health, Johns Hopkins Bloomberg School of Public Health, Baltimore, USA; Department of Nursing, Temple University College of Public Health, Philadelphia, USA; Department of Epidemiology, Johns Hopkins Bloomberg School of Public Health, Baltimore, USA

## Abstract

Diarrheal illnesses remain a leading cause of morbidity and mortality worldwide. Understanding when and where individuals seek healthcare is essential for accurately assessing disease burden and improving access to appropriate care. In this study, we conducted a meta-synthesis of qualitative research examining barriers and facilitators to care-seeking for diarrheal illness, either for individuals themselves or for their children. Specifically, we performed a systematic review of qualitative studies from any geographic location and time period that explored motivations for seeking care outside the home for diarrheal illness across all age groups. We then conducted a thematic analysis of the included studies to identify recurring patterns related to care-seeking behavior and to synthesize qualitative insights across contexts. In total, 47 studies met our inclusion criteria, the majority of which focused on caregiver decision-making for children with diarrhea in sub-Saharan Africa. Our meta-synthesis identified several key factors that influenced whether and when individuals sought care. Sociocultural norms, including locally held beliefs about disease causation, were frequently cited as influencing decisions to seek or delay formal care. Additional barriers included long travel distances to health facilities, transportation costs, limited trust in healthcare providers, negative feelings, and inconsistent availability of care. Conversely, episodes perceived as severe were more likely to prompt care-seeking outside the home. These findings highlight the importance of contextually grounded interventions that improve physical and financial access to care, foster trust in healthcare providers through consistent and effective service delivery and strengthen community engagement around recognizing signs of severe illness and the potential benefits of timely treatment. They also underscore the need for future studies to define diarrhea in locally relevant terms and to clearly define sources of care-seeking, as variation in these definitions can limit our full understanding of who is affected and how individuals respond to illness.

## Introduction

Diarrheal diseases were associated with an estimated 4.7 billion incident cases and 1.2 million deaths in 2021 [1]. Many of these cases and deaths could have been averted through access to clean water, vaccination, proper nutrition, and appropriate case management and treatment [2,3]. Appropriate diarrheal treatment may involve rehydration, either intravenously or orally through oral rehydration solutions, zinc supplementation, and/or use of antibiotics [4]. While oral diarrhea treatments are effective and generally inexpensive, they often require seeking care outside the home at a health post, pharmacy, hospital, or clinic. Care-seeking at a public hospital or clinic is also required for cases to be reported to health systems and counted in estimates of diarrhea burden, which are used to distribute limited diarrhea mitigation resources such as vaccines. Thus, understanding care-seeking behavior for diarrhea is important for improving access to treatment, estimating diarrhea burden, and preventing cases and deaths.

To better understand how many community diarrhea cases go unreported and possibly untreated, we previously conducted a systematic review and meta-analysis of studies that reported the proportion of individuals or caregivers that seek care when they or their child had diarrheal illness [5]. We found substantial variation across the 166 quantitative studies identified for inclusion in the proportion of people who sought medical care, with estimates ranging from nearly 0% to 100% depending on the study context. The only factor that was consistently associated with this variation in care-seeking was diarrhea case definition, with care-seeking significantly higher for severe diarrhea or cholera compared to general diarrhea. However, given wide variation in the data that was reported in the 166 studies, we were not able to identify other common reasons that people do or do not seek care.

Secondary analyses of individual-level care-seeking survey data have found that education, healthcare knowledge and experience, wealth, gender roles, and autonomy in decision making are all associated with variation in care-seeking for diarrheal illness [6–10]. A limitation of these studies and the systematic review is that they used survey data collected using pre-specified questionnaires and did not have the flexibility to examine nuanced reasons that individuals do or do not seek care. This limits our ability to understand what can be done to improve access to care, especially for hard-to-reach populations that are systematically not captured by surveillance due to low interaction with the health care system. Qualitative studies that employ open-ended questionnaires and semi-structured interviews provide an opportunity to address this gap.

In this study, we synthesized information from qualitative studies to identify common themes and reasons that individuals or caregivers do not seek care at medical facilities when they or their child have diarrhea. We identified common barriers and motivations to care-seeking across studies, as well as barriers specific to study settings. The findings may help to develop interventions that lower barriers to seeking care and/or identify alternate methods of collecting surveillance data and delivering care, by leveraging insights into patterns of care-seeking to collect data and engage with informal care settings.

## Methods

We conducted a qualitative meta-synthesis using a rigorous methodological approach to identify broad themes and insights across multiple study settings. Previous studies have demonstrated that this process can lead to a comprehensive understanding of complex issues, which is especially beneficial for interpreting complex care-seeking behaviors [11–14].

### Search Strategy

Mixed-methods and qualitative studies were identified by a previous systematic review of care-seeking for diarrhea [15]. Briefly, PubMed, Embase, Web of Science, and Global Index Medicus were initially searched on January 23, 2023, and the search was updated on September 3, 2024. The review included quantitative and qualitative studies examining the proportion of people with a diarrheal illness that sought care for themselves or their child. Any study reporting mixed or qualitative methods during title and abstract or full text screening was flagged for inclusion in this qualitative meta-synthesis [15]. Exact search terms and further descriptions of study methods can be found in the manuscript and supplementary materials of Wiens et al. [15].

### Eligibility Criteria

We included studies that employed a qualitative study design to examine reasons individuals did or did not seek care for themselves or a child with diarrheal illness from any location and time period. We did not set limits on language, publication date, or sample size. We excluded studies that 1) did not report care-seeking motivations specifically for diarrhea (e.g., discussed general childhood illnesses), 2) did not report data obtained through qualitative study design methods, including individual interviews, focus group discussions, or other forms of open-ended study design, 3) were reviews or secondary analyses, or 4) did not have full texts available.

### Study Screening Methods

Identified studies were uploaded to Covidence (https://covidence.org) and screened for eligibility. Full texts were assessed for eligibility by two independent reviewers (MHM, SMH, ENR, KEW). A third reviewer (MHM, SMH, ENR, KEW) served as a tie breaking vote when needed.

### Data Extraction and Coding

Two reviewers (MHM, SMH) extracted study information from included studies in table format. All included variables and their definitions can be found in S1 Data. All study variables were recorded when reported, but none were required. As this was a scoping review, we did not assess the quality of the qualitative data. While the Critical Appraisal Skills Programme [CASP]’s checklist for evaluating the quality of qualitative studies exists [16], there is no agreement on what score merits exclusion [17,18].

Two reviewers (MHM, SMH) coded factors influencing care-seeking decisions using Atlas.ti Web (https://atlasti.com/atlas-ti-web). The two reviewers divided the articles to code, extracted qualitative data, and then reviewed each other’s work to assess agreement. Throughout this process both reviewers kept audit journals, tracking any questions, comments or thoughts they had during this process to ensure transparency and reproducibility, and reduce bias. For the updated search, a single reviewer (MHM) extracted and qualitatively coded data in accordance with the parent study [15]. A third reviewer (KEW, ENR) moderated group reconciliation meetings to resolve any disagreements, and all reviewers agreed on the final decision.

We developed the codebook using a structured approach informed by thematic analysis and grounded theory principles. We began the process by brainstorming potential influences on care-seeking behaviors informed by the research team’s existing understanding, as well as concepts the full text reviewers identified during the screening process. A single reviewer (ENR) then conducted a preliminary screening of a small sample of included, geographically distinct studies using open coding. Subsequently, we combined the concepts already identified as potentially relevant with any new concepts that emerged from the screening and open coding to create a preliminary codebook. We structured the codebook hierarchically, with parent codes representing anticipated major themes and child-codes capturing more specific aspects of each theme. As we worked through this process, we continuously compared our codes to ensure each one was clearly defined and did not overlap with others.

We applied the codebook to all included studies and refined it as needed when information in the articles did not clearly fit within the existing codes. For example, we expanded the “external other” code to also capture religious ideas. Similar ideas were managed as groups under one parent code in Atlas.ti, with individual child codes in the parent group used for thematic synthesis. The final codebook used by reviewers is shown in Table S1.

### Thematic Analysis

After coding all studies, we summarized and reviewed the content included under each child code to search for broader patterns of meaning. Within each code, we identified themes to capture the nuances of the data and structure the main themes. To develop a comprehensive thematic framework, two researchers (MHM, SHM) independently created model theme trees based on patterns in the coded data. We then presented these frameworks to the full research team, using their input to refine the two frameworks into a single structure. After this initial restructuring, we conducted two rounds of feedback to ensure all themes and sub-themes were comprehensive, distinct from each other, and answered the research question.

## Results

### Search Results

After screening of 5,188 studies, Wiens et al. identified 106 studies for review [15]. Following full text screening, 47 studies met the inclusion criteria for our meta-synthesis (Fig 1).

**Fig 1.**
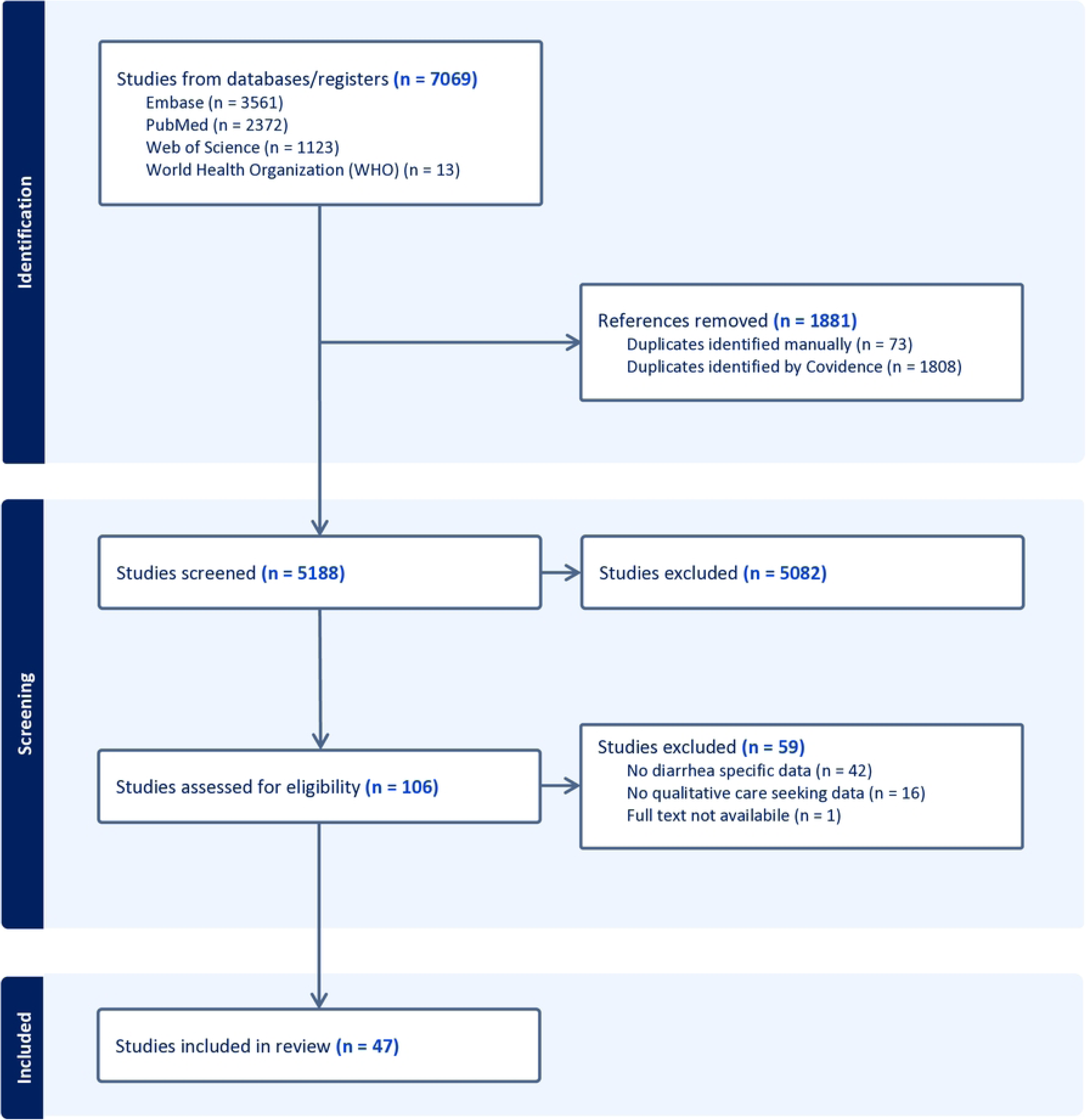
Prisma diagram. Preferred reporting items for systematic reviews and meta-analyses diagram of search and inclusion. Diagram shows the search, screening, and inclusion process for the systematic review, including the databases searched and reasons for studies were excluded. Studies were identified and initially screened in a previous review [15].

### Characteristics of Included Studies

The included studies are detailed in Table 1; additional study information can be found in S1 Data. The studies came from 25 countries, with most from Sub-Saharan Africa (n = 23) and Southeast Asia (n = 15). The remaining studies were from North and South America (n = 6), the Middle East (n = 2), and Oceania (n = 1). Twenty-five studies were conducted in rural areas and ten in urban areas, with the remaining twelve study locations in mixed rural-urban settings. Among studies with information about sampling dates (n = 39), completion dates ranged from 1993 to 2021, with the majority from 2010 to 2014 (n = 14). Studies employed individual interviews (n = 18), group interviews (n = 5), or a mix of both (n = 24). Most studies asked about care-seeking practices for children with diarrhea (n = 39), but some asked about care-seeking for the respondents themselves (n = 9) and/or a hypothetical diarrheal episode (n = 4). Thirty-five studies did not provide an explicit definition for diarrhea, while the remainder used the World Health Organization’s case definition ([19]) (n=3), acute gastroenteritis (n = 4), or cholera/severe diarrhea (n=5).

**Table 1.**
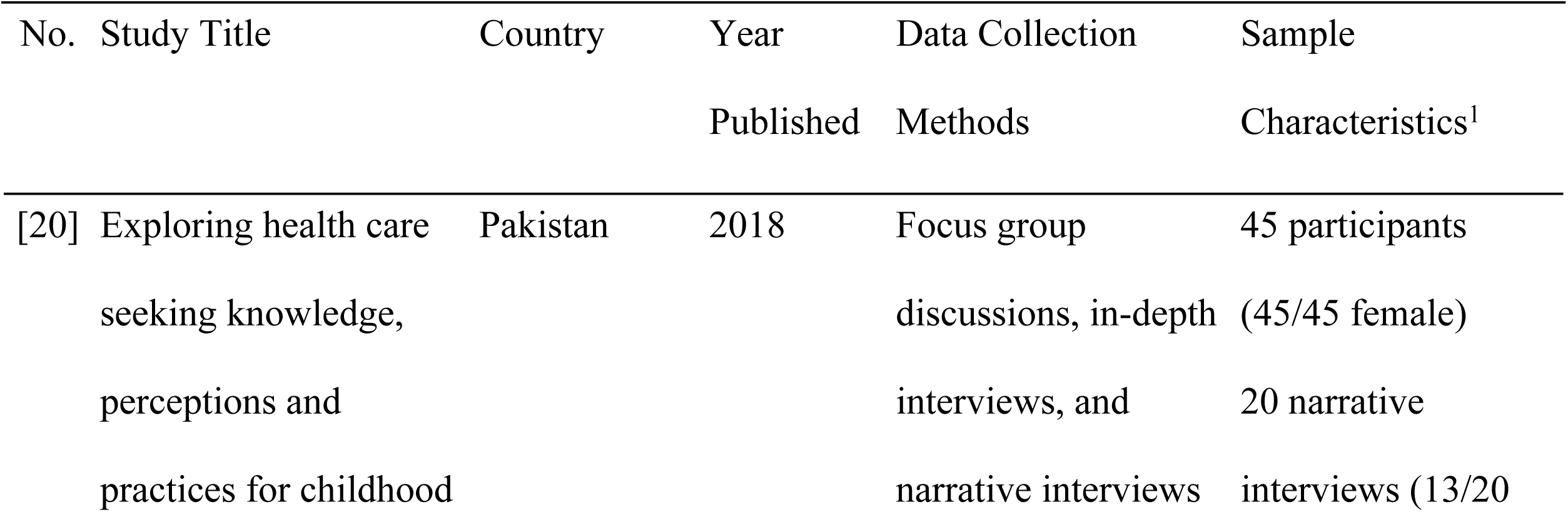
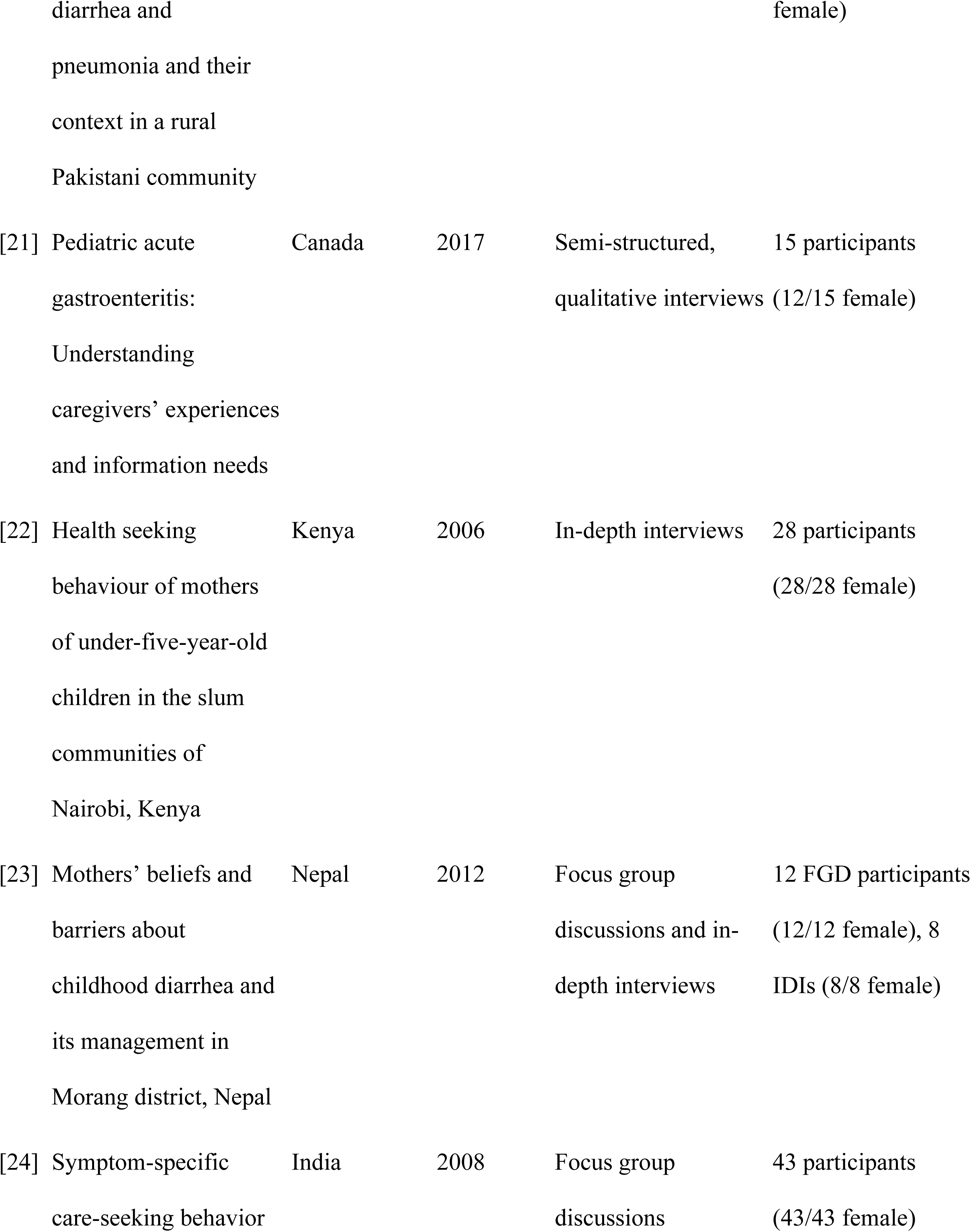
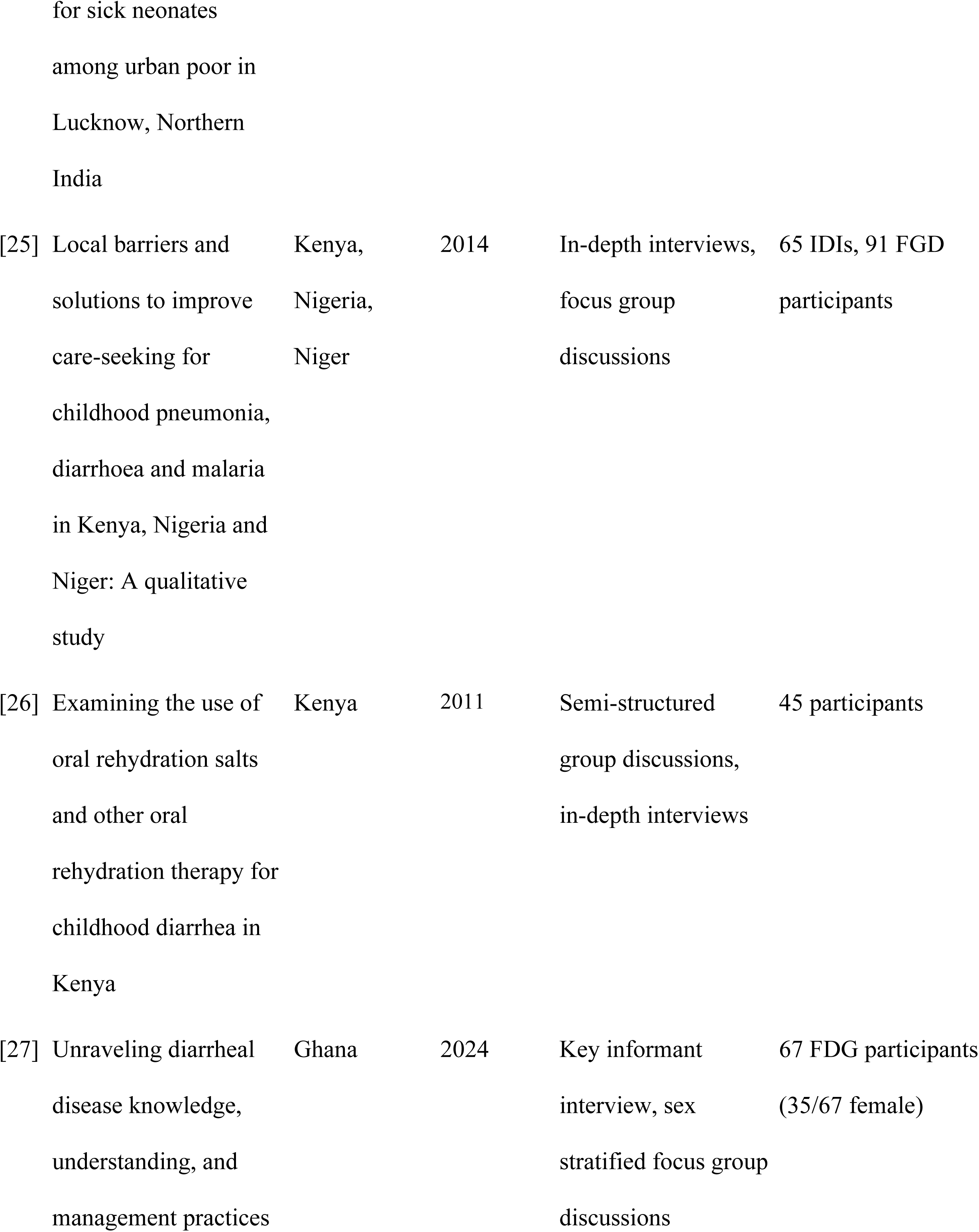
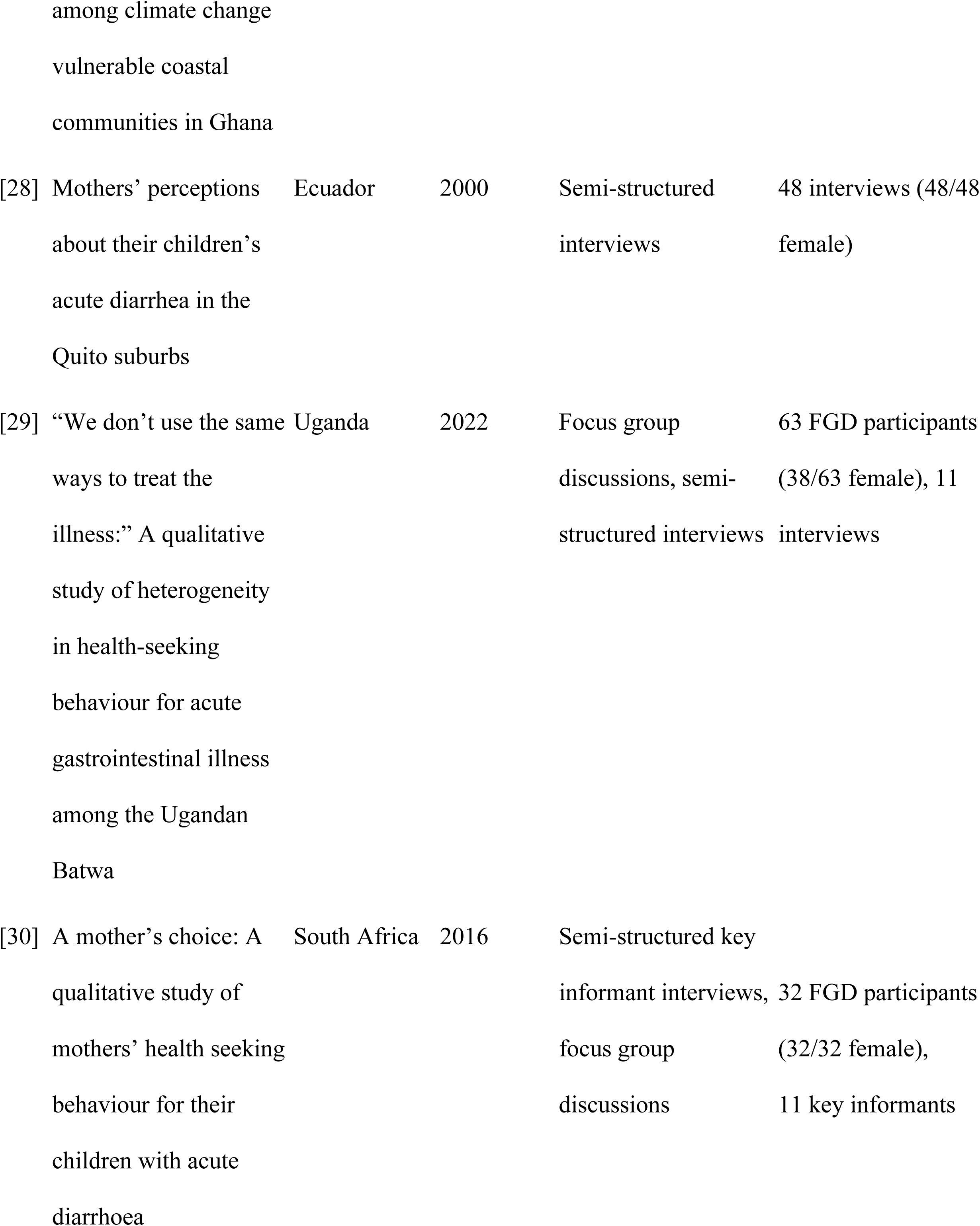
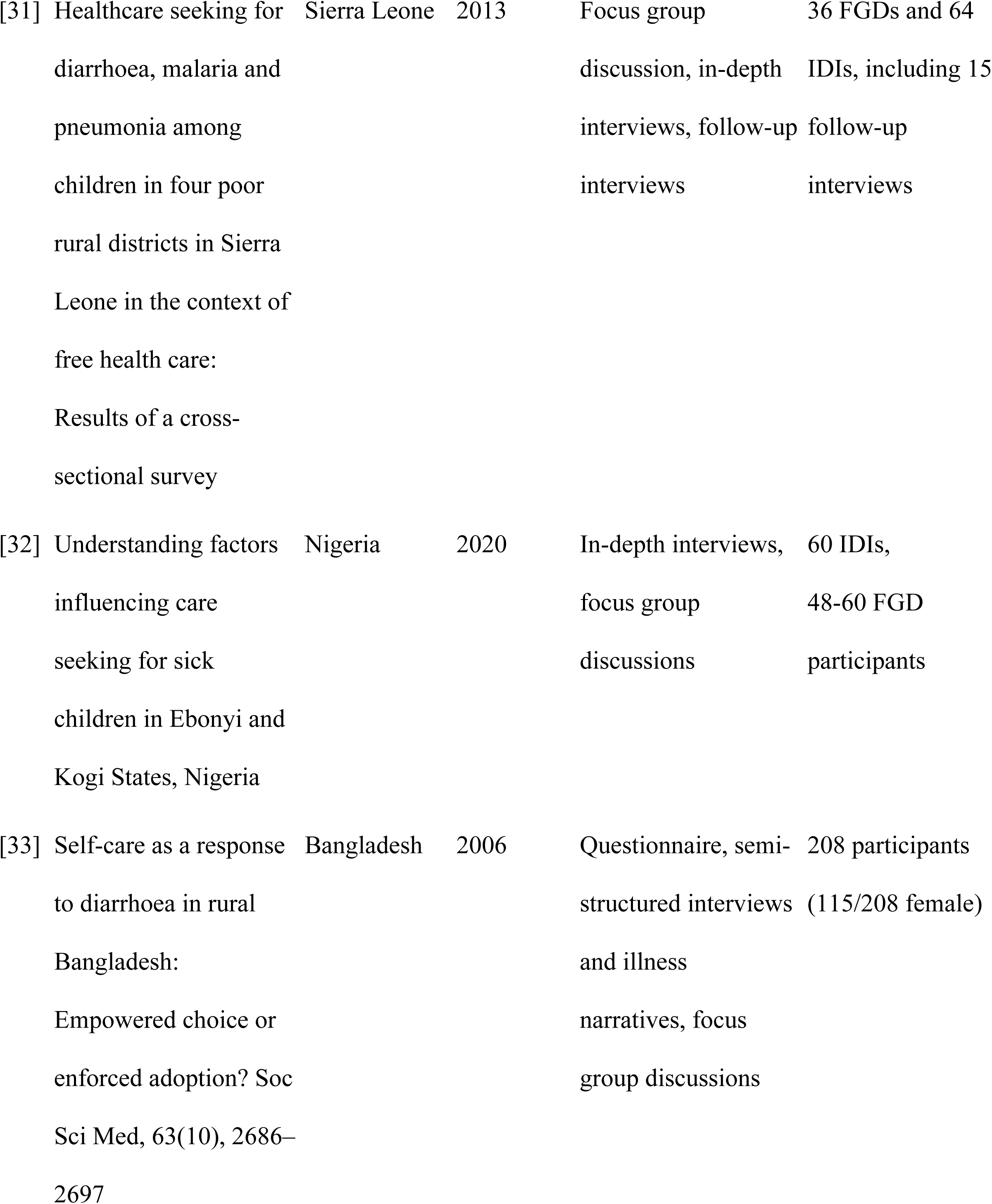
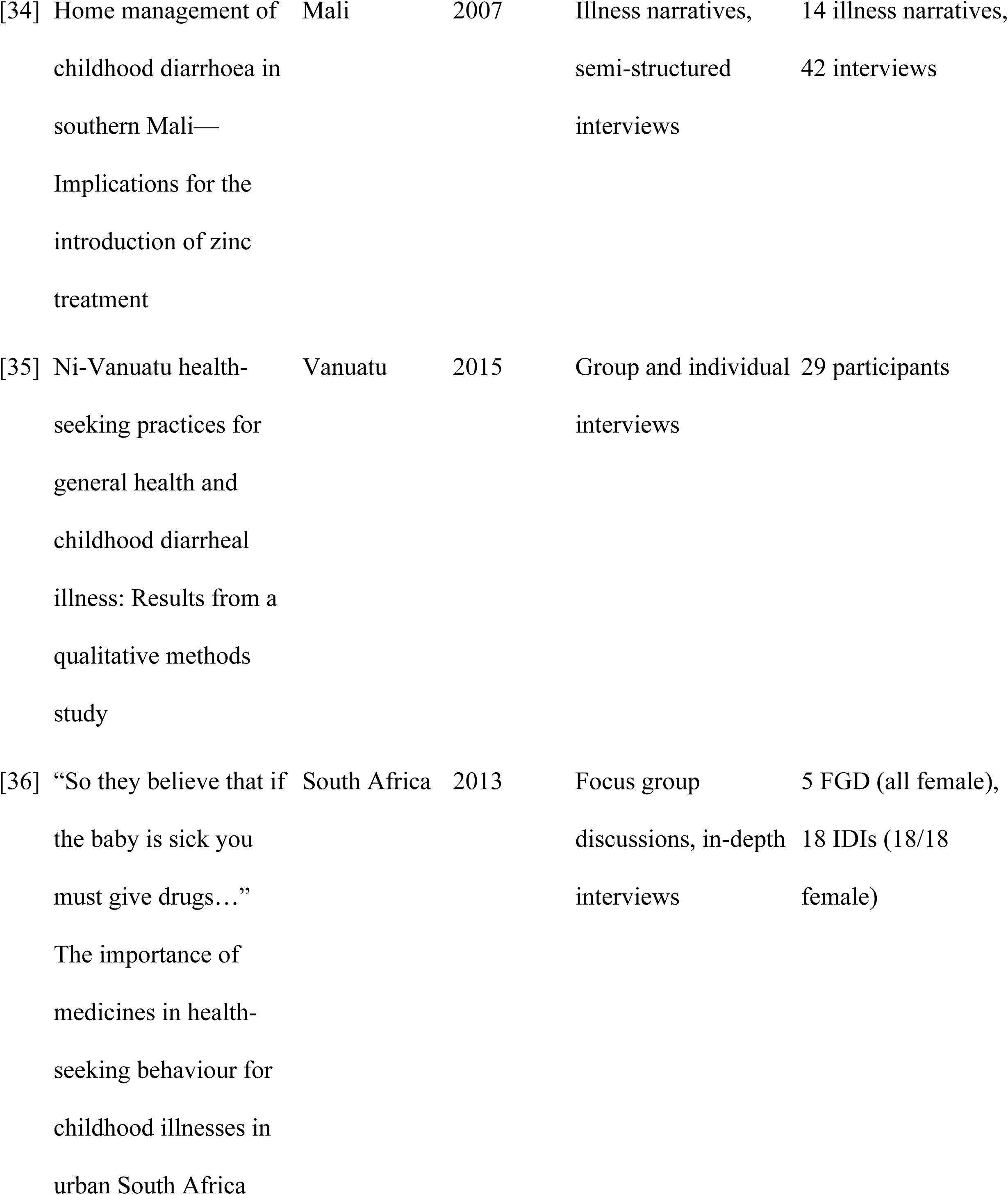
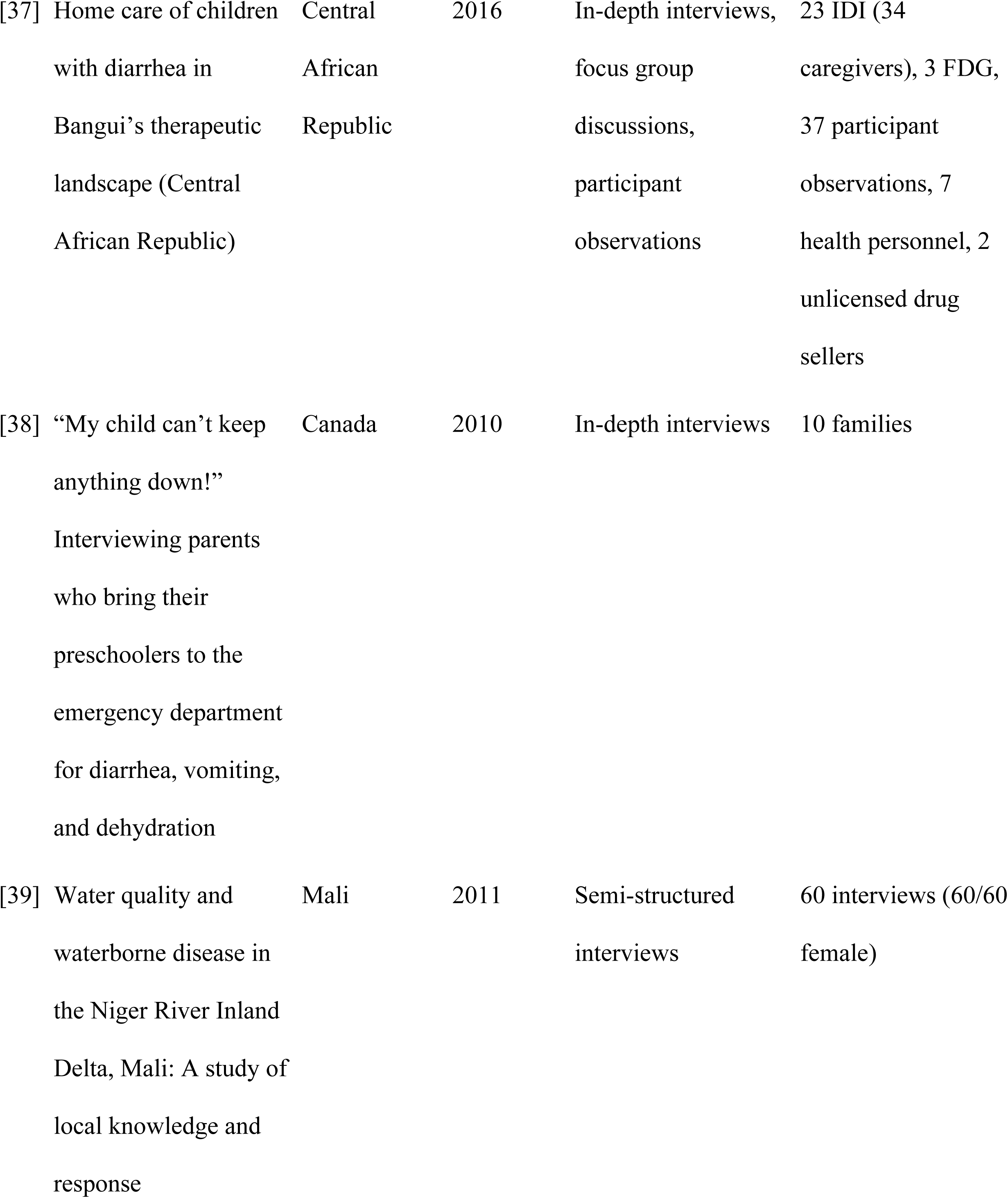
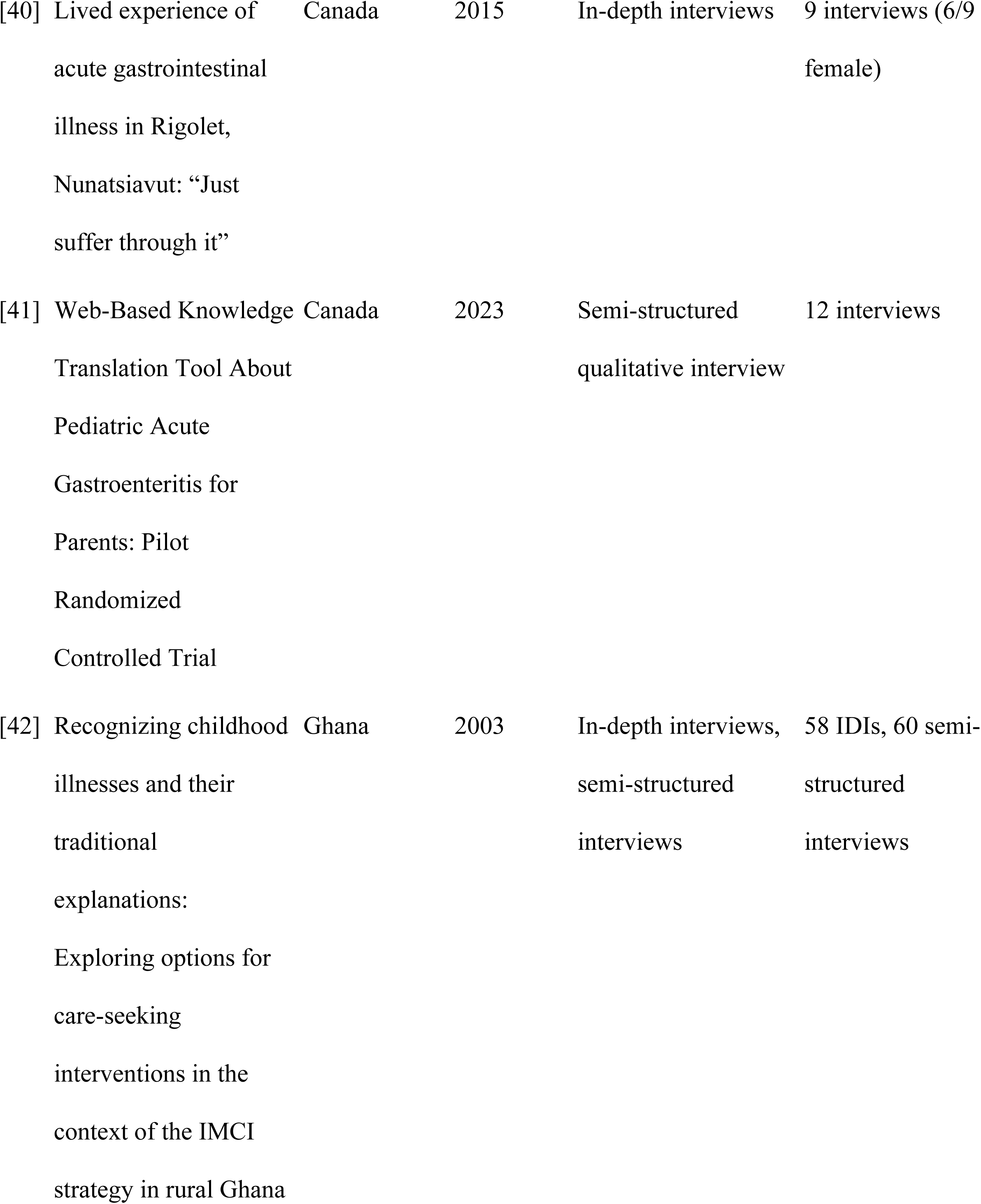
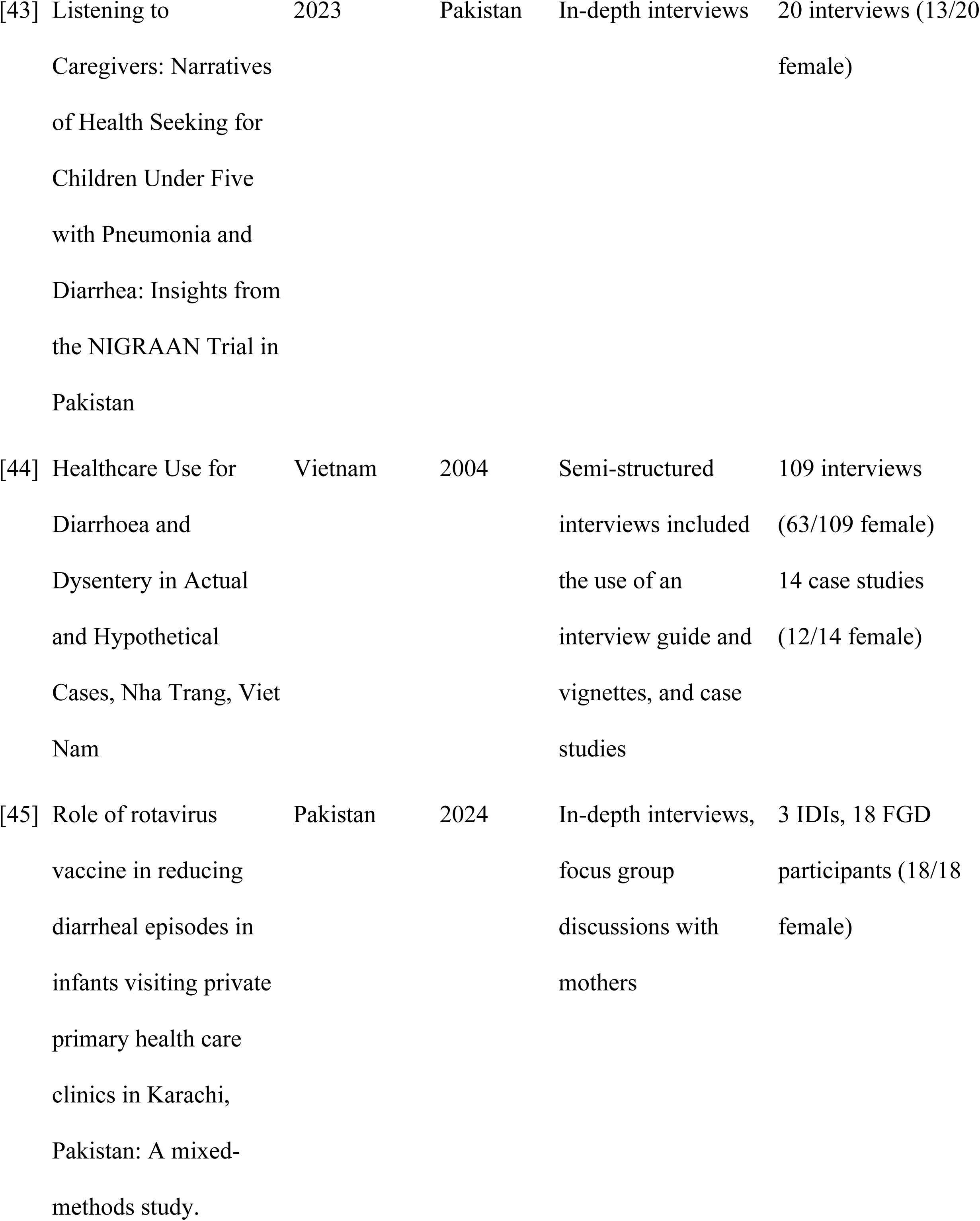
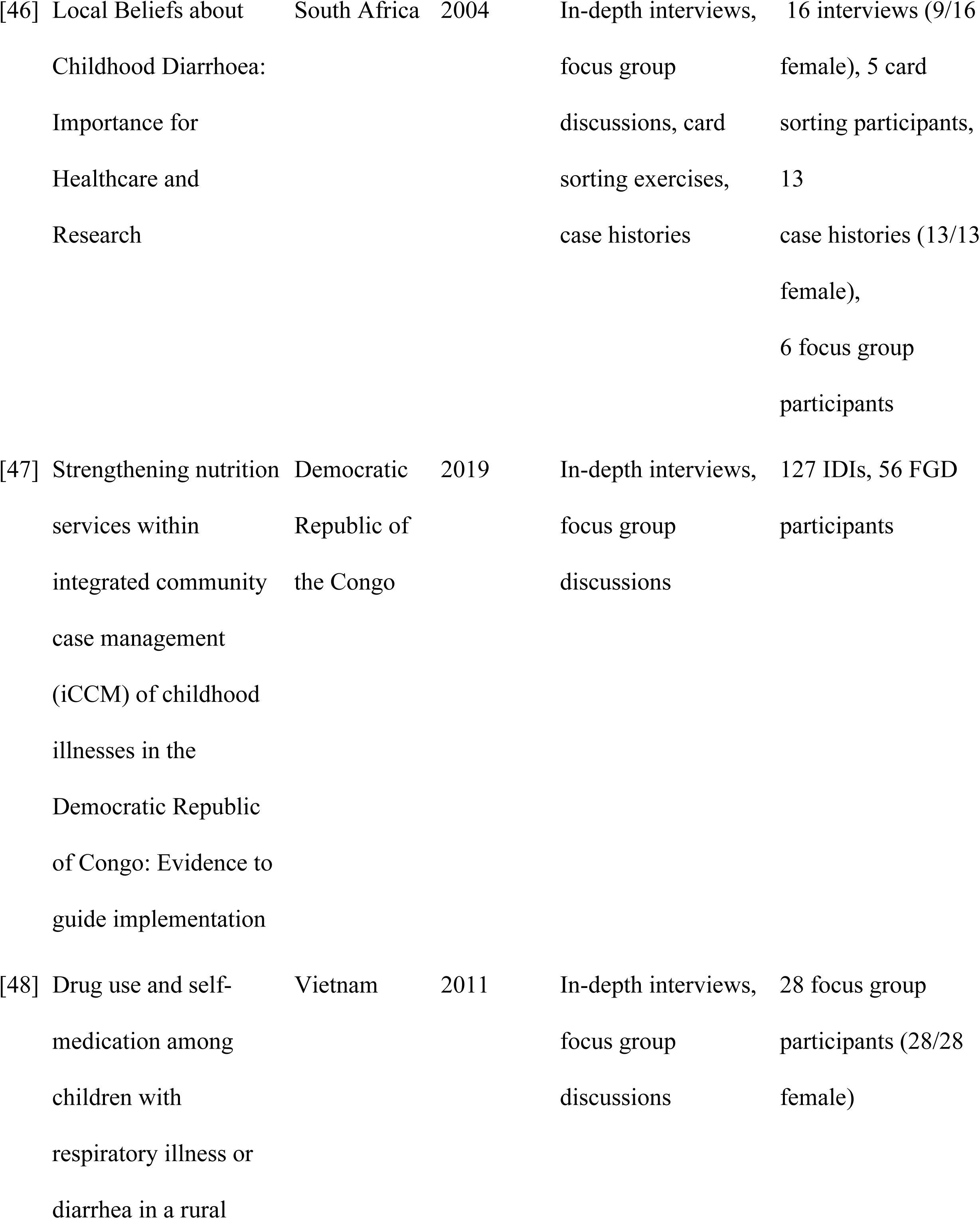
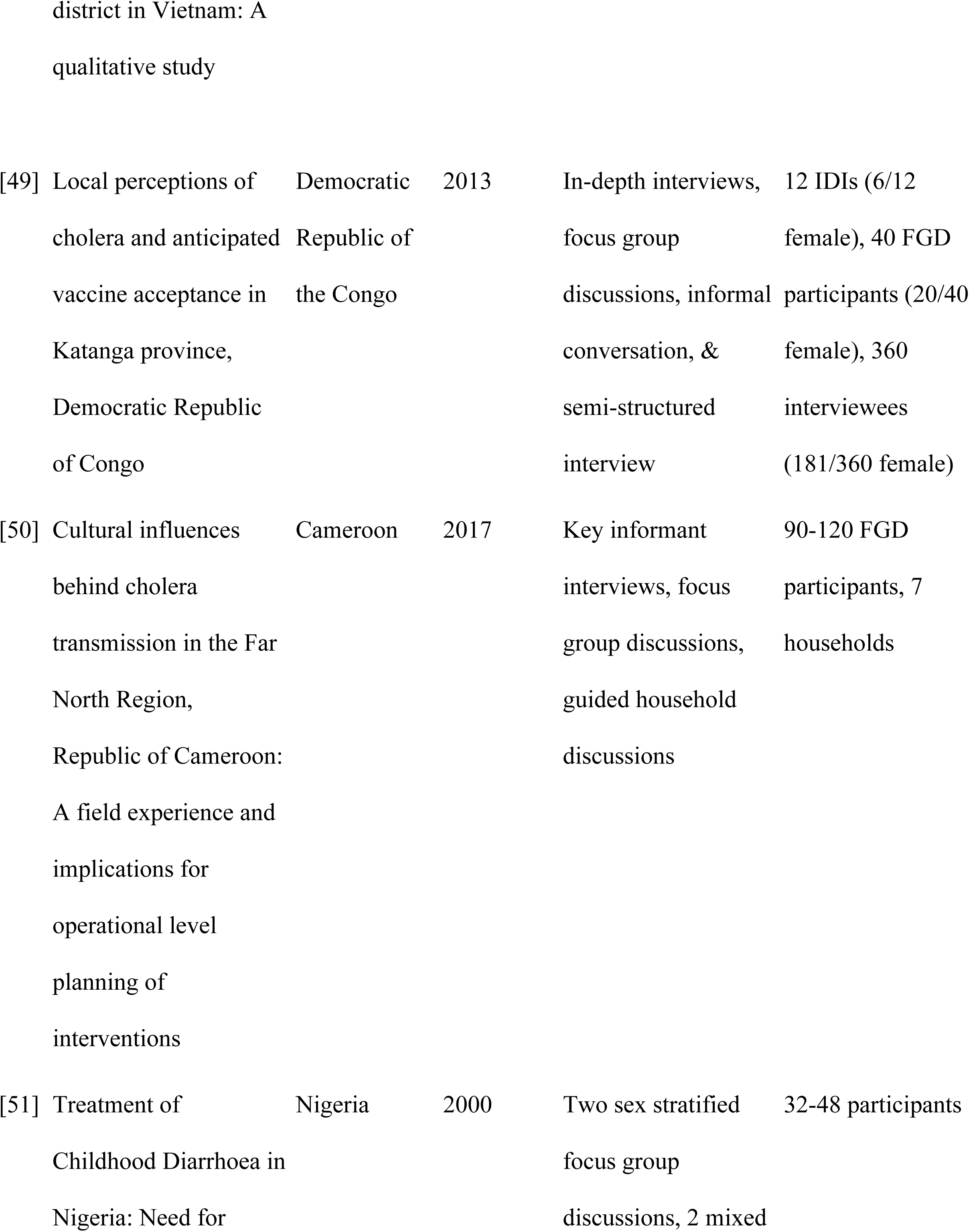
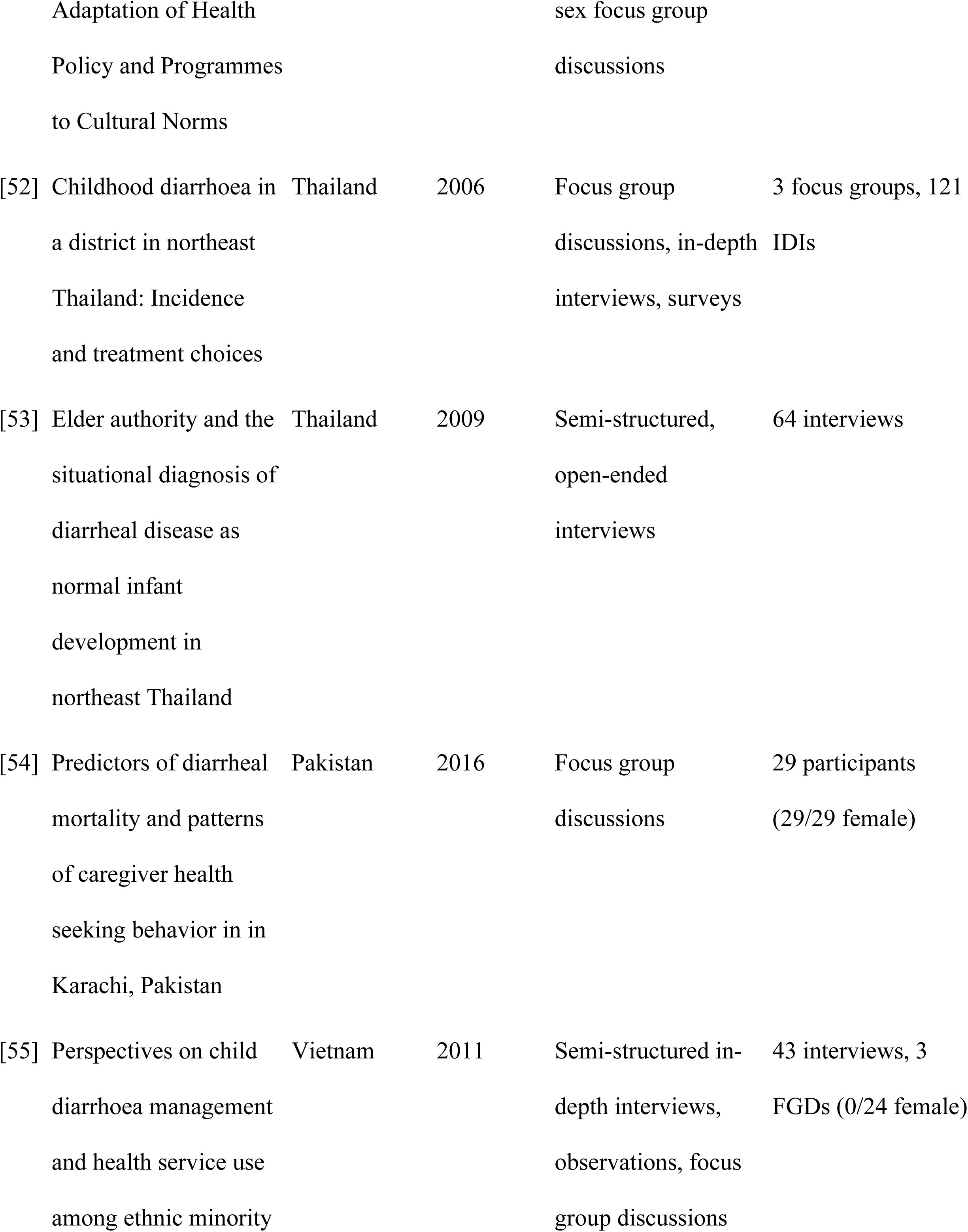
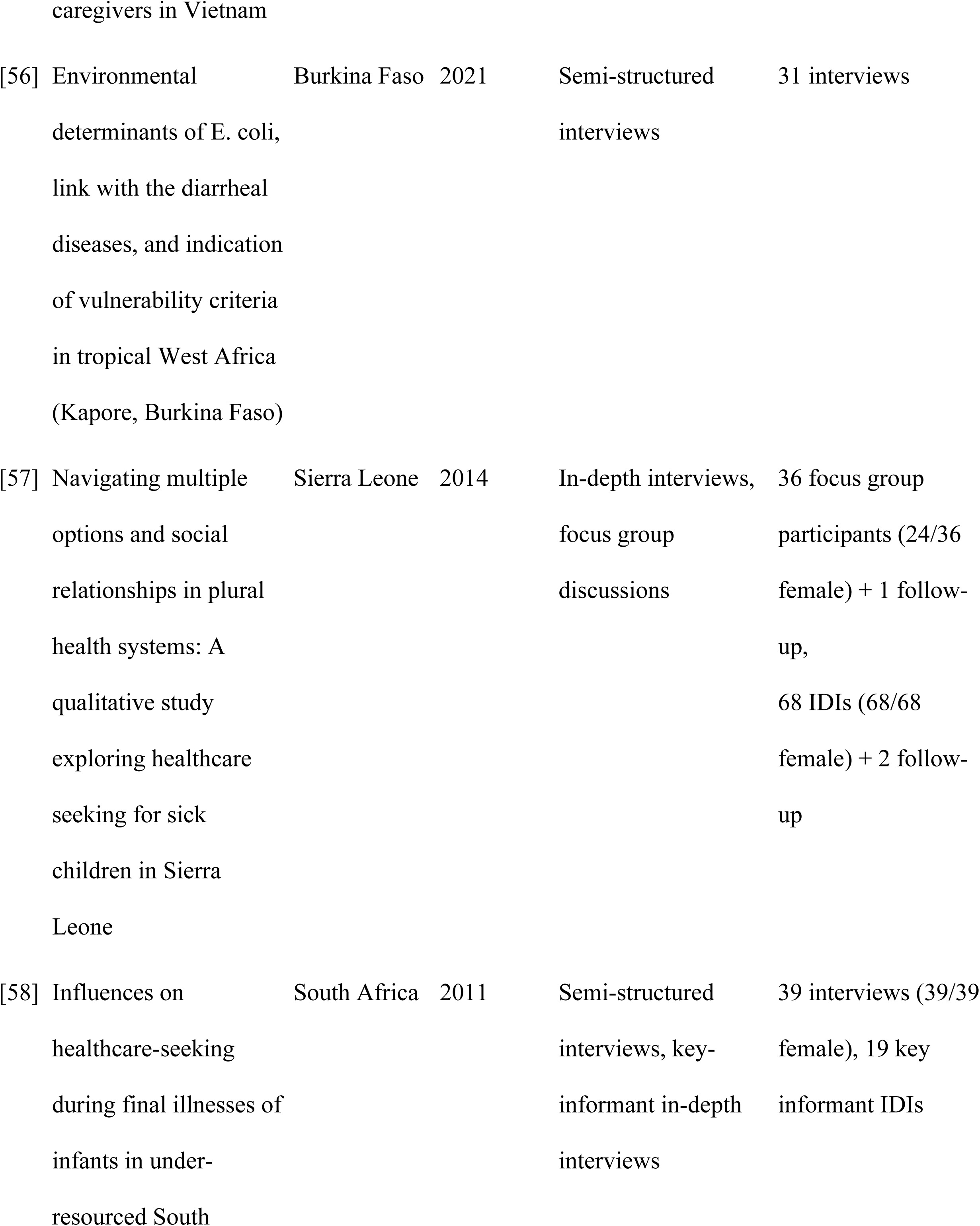
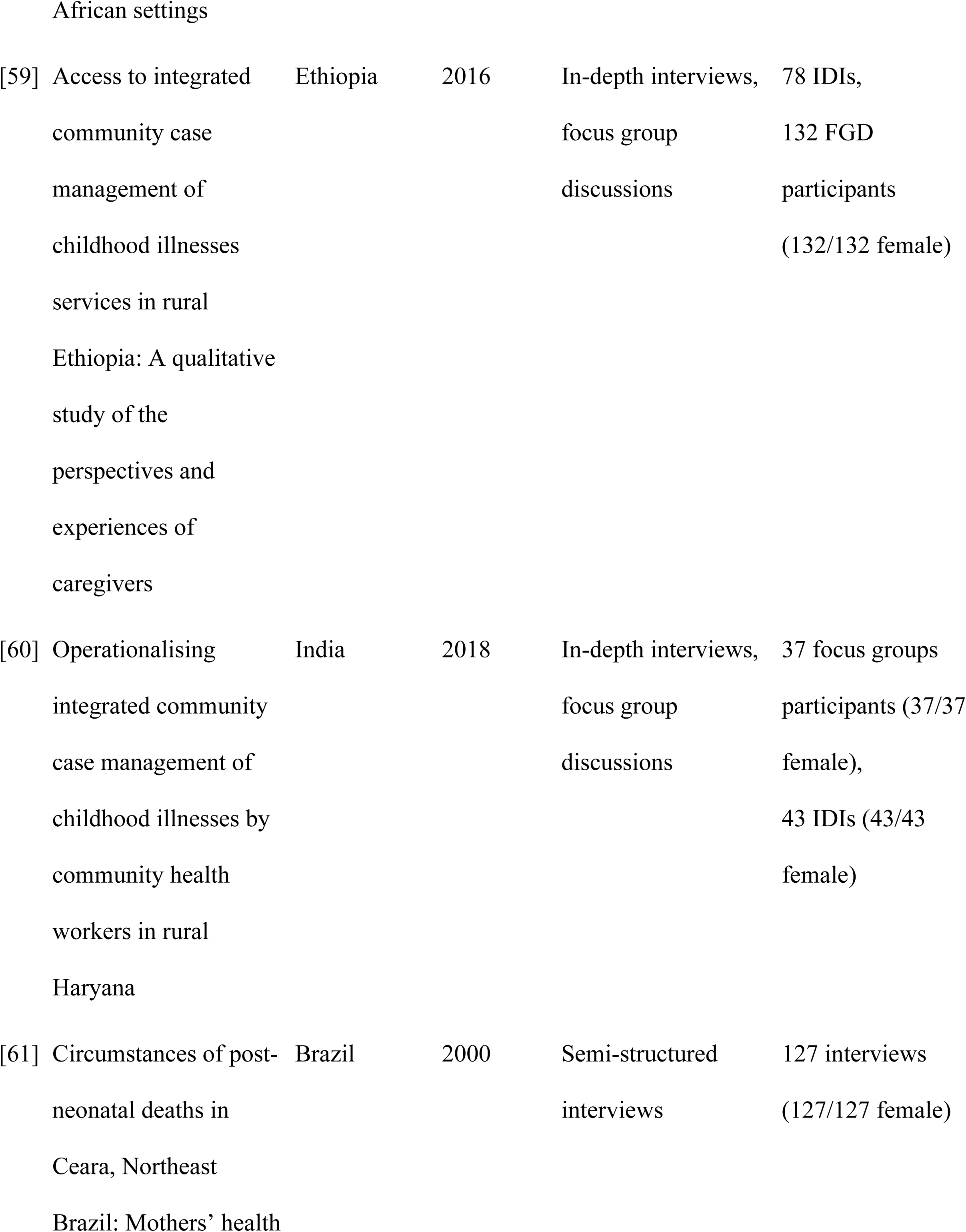
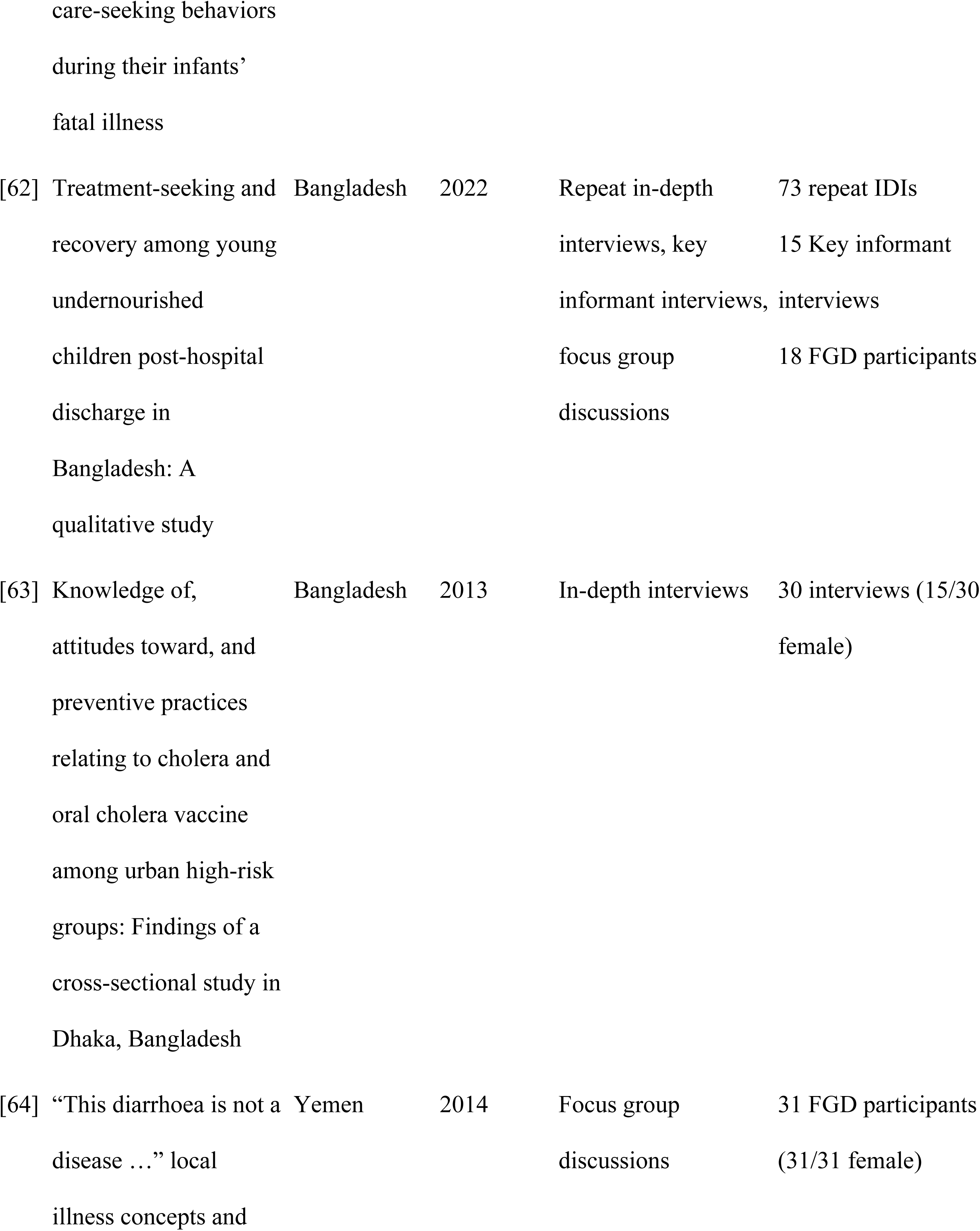
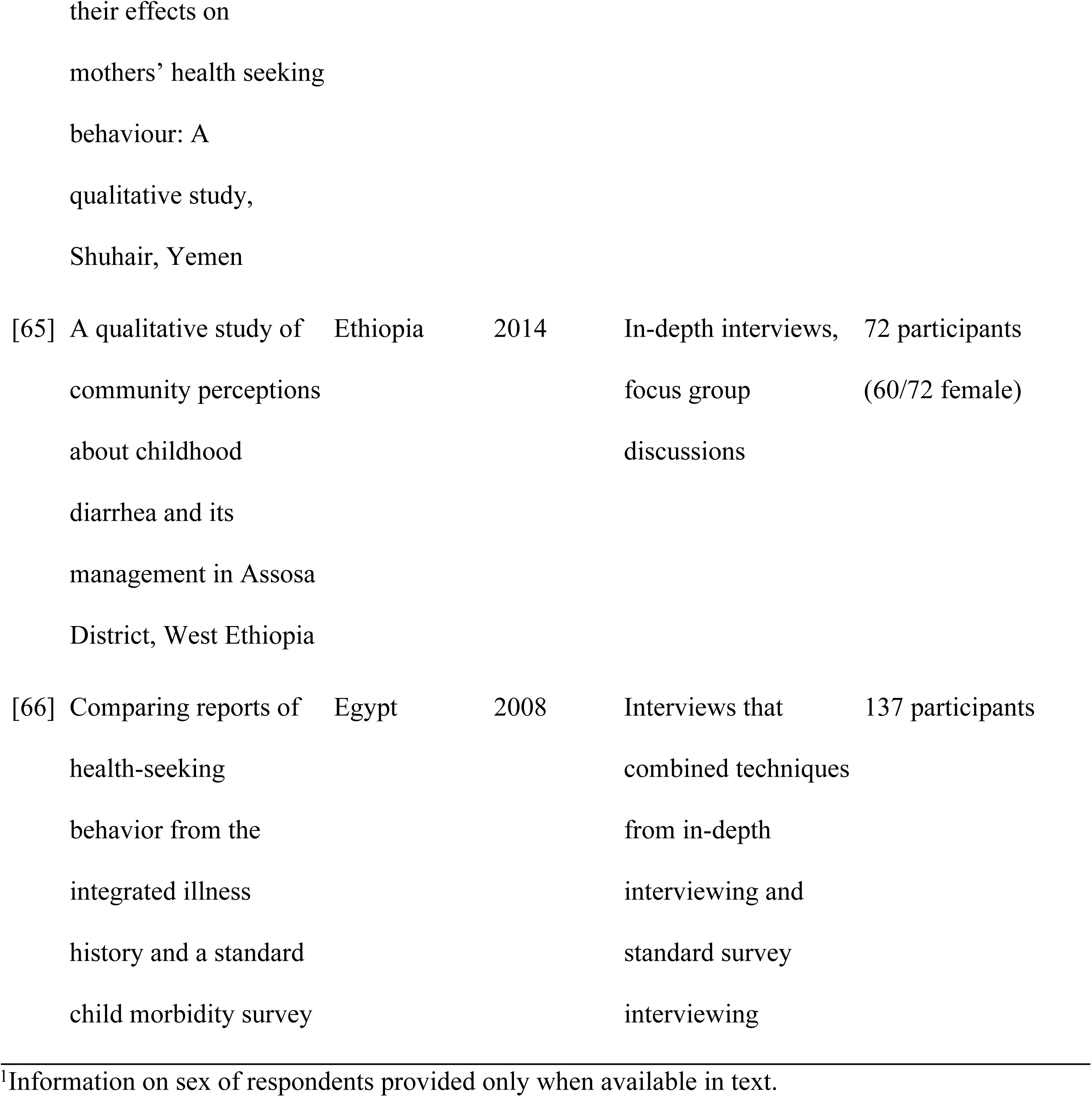
Summary of included studies.

| No. | Study Title | Country | Year<br>Published | Data Collection |  | Sample<br>Characteristics<sup>1</sup> |
| --- | --- | --- | --- | --- | --- | --- |
|  |  |  |  | Methods |  |  |
| [20] | Exploring health care seeking knowledge, perceptions and practices for childhood | Pakistan | 2018 | Focus group discussions, in-depth interviews, and narrative interviews |  | 45 participants (45/45 female)<br>20 narrative interviews (13/20 |

|  |  |  |  |  |  |
| --- | --- | --- | --- | --- | --- |
|  | diarrhea and |  |  |  | female) |
|  | pneumonia and their |  |  |  |  |
|  | context in a rural |  |  |  |  |
|  | Pakistani community |  |  |  |  |
| [21] | Pediatric acute gastroenteritis: Understanding caregivers' experiences and information needs | Canada | 2017 | Semi-structured, qualitative interviews | 15 participants (12/15 female) |
| [22] | Health seeking behaviour of mothers of under-five-year-old children in the slum communities of Nairobi, Kenya | Kenya | 2006 | In-depth interviews | 28 participants (28/28 female) |
| [23] | Mothers' beliefs and barriers about childhood diarrhea and its management in Morang district, Nepal | Nepal | 2012 | Focus group discussions and in-depth interviews | 12 FGD participants (12/12 female), 8 IDIs (8/8 female) |
| [24] | Symptom-specific care-seeking behavior | India | 2008 | Focus group discussions | 43 participants (43/43 female) |

|  |  |  |  |  |  |
| --- | --- | --- | --- | --- | --- |
| [25] | Local barriers and solutions to improve care-seeking for childhood pneumonia, diarrhoea and malaria in Kenya, Nigeria and Niger: A qualitative study | Kenya, Nigeria, Niger | 2014 | In-depth interviews, focus group discussions | 65 IDIs, 91 FGD participants |
| [26] | Examining the use of oral rehydration salts and other oral rehydration therapy for childhood diarrhea in Kenya | Kenya | 2011 | Semi-structured group discussions, in-depth interviews | 45 participants |
| [27] | Unraveling diarrheal disease knowledge, understanding, and management practices | Ghana | 2024 | Key informant interview, sex stratified focus group discussions | 67 FDG participants (35/67 female) |

|  |  |  |  |  |  |
| --- | --- | --- | --- | --- | --- |
| [28] | Mothers' perceptions about their children's acute diarrhea in the Quito suburbs | Ecuador | 2000 | Semi-structured interviews | 48 interviews (48/48 female) |
| [29] | “We don't use the same ways to treat the illness.” A qualitative study of heterogeneity in health-seeking behaviour for acute gastrointestinal illness among the Ugandan Batwa | Uganda | 2022 | Focus group discussions, semi-structured interviews | 63 FGD participants (38/63 female), 11 interviews |
| [30] | A mother's choice: A qualitative study of mothers' health seeking behaviour for their children with acute diarrhoea | South Africa | 2016 | Semi-structured key informant interviews, focus group discussions | 32 FGD participants (32/32 female), 11 key informants |
| [31] | Healthcare seeking for diarrhoea, malaria and pneumonia among children in four poor rural districts in Sierra Leone in the context of free health care: Results of a cross-sectional survey | Sierra Leone | 2013 | Focus group discussion, in-depth interviews, follow-up interviews | 36 FGDs and 64 IDIs, including 15 follow-up interviews |
| [32] | Understanding factors influencing care seeking for sick children in Ebonyi and Kogi States, Nigeria | Nigeria | 2020 | In-depth interviews, focus group discussions | 60 IDIs, 48-60 FGD participants |
| [33] | Self-care as a response to diarrhoea in rural Bangladesh: Empowered choice or enforced adoption? Soc Sci Med, 63(10), 2686–2697 | Bangladesh | 2006 | Questionnaire, semi-structured interviews and illness narratives, focus group discussions | 208 participants (115/208 female) |
| [34] | Home management of childhood diarrhoea in southern Mali—<br>Implications for the introduction of zinc treatment | Mali | 2007 | Illness narratives, semi-structured interviews | 14 illness narratives, 42 interviews |
| [35] | Ni-Vanuatu health-seeking practices for general health and childhood diarrheal illness: Results from a qualitative methods study | Vanuatu | 2015 | Group and individual interviews | 29 participants |
| [36] | “So they believe that if the baby is sick you must give drugs...”<br>The importance of medicines in health-seeking behaviour for childhood illnesses in urban South Africa | South Africa | 2013 | Focus group discussions, in-depth interviews | 5 FGD (all female), 18 IDIs (18/18 female) |
| [37] | Home care of children with diarrhea in Bangui's therapeutic landscape (Central African Republic) | Central African Republic | 2016 | In-depth interviews, focus group discussions, participant observations | 23 IDI (34 caregivers), 3 FDG, 37 participant observations, 7 health personnel, 2 unlicensed drug sellers |
| [38] | "My child can't keep anything down!"<br>Interviewing parents who bring their preschoolers to the emergency department for diarrhea, vomiting, and dehydration | Canada | 2010 | In-depth interviews | 10 families |
| [39] | Water quality and waterborne disease in the Niger River Inland Delta, Mali: A study of local knowledge and response | Mali | 2011 | Semi-structured interviews | 60 interviews (60/60 female) |
| [40] | Lived experience of acute gastrointestinal illness in Rigolet, Nunatsiavut: “Just suffer through it” | Canada | 2015 | In-depth interviews | 9 interviews (6/9 female) |
| [41] | Web-Based Knowledge Translation Tool About Pediatric Acute Gastroenteritis for Parents: Pilot Randomized Controlled Trial | Canada | 2023 | Semi-structured qualitative interview | 12 interviews |
| [42] | Recognizing childhood illnesses and their traditional explanations: Exploring options for care-seeking interventions in the context of the IMCI strategy in rural Ghana | Ghana | 2003 | In-depth interviews, semi-structured interviews | 58 IDIs, 60 semi-structured interviews |
| [43] | Listening to<br>Caregivers: Narratives<br>of Health Seeking for<br>Children Under Five<br>with Pneumonia and<br>Diarrhea: Insights from<br>the NIGRAAN Trial in<br>Pakistan | 2023 | Pakistan | In-depth interviews | 20 interviews (13/20<br>female) |
| [44] | Healthcare Use for<br>Diarrhoea and<br>Dysentery in Actual<br>and Hypothetical<br>Cases, Nha Trang, Viet<br>Nam | Vietnam | 2004 | Semi-structured<br>interviews included<br>the use of an<br>interview guide and<br>vignettes, and case<br>studies | 109 interviews<br>(63/109 female)<br>14 case studies<br>(12/14 female) |
| [45] | Role of rotavirus<br>vaccine in reducing<br>diarrheal episodes in<br>infants visiting private<br>primary health care<br>clinics in Karachi,<br>Pakistan: A mixed-methods study. | Pakistan | 2024 | In-depth interviews,<br>focus group<br>discussions with<br>mothers | 3 IDIs, 18 FGD<br>participants (18/18<br>female) |
| [46] | Local Beliefs about Childhood Diarrhoea: Importance for Healthcare and Research | South Africa | 2004 | In-depth interviews, focus group discussions, card sorting exercises, case histories | 16 interviews (9/16 female), 5 card sorting participants, 13 case histories (13/13 female), 6 focus group participants |
| [47] | Strengthening nutrition services within integrated community case management (iCCM) of childhood illnesses in the Democratic Republic of Congo: Evidence to guide implementation | Democratic Republic of the Congo | 2019 | In-depth interviews, focus group discussions | 127 IDIs, 56 FGD participants |
| [48] | Drug use and self-medication among children with respiratory illness or diarrhea in a rural | Vietnam | 2011 | In-depth interviews, focus group discussions | 28 focus group participants (28/28 female) |

|  |  |  |  |  |  |
| --- | --- | --- | --- | --- | --- |
| [49] | Local perceptions of cholera and anticipated vaccine acceptance in Katanga province, Democratic Republic of Congo | Democratic Republic of the Congo | 2013 | In-depth interviews, focus group discussions, informal conversation, & semi-structured interview | 12 IDIs (6/12 female), 40 FGD participants (20/40 female), 360 interviewees (181/360 female) |
| [50] | Cultural influences behind cholera transmission in the Far North Region, Republic of Cameroon: A field experience and implications for operational level planning of interventions | Cameroon | 2017 | Key informant interviews, focus group discussions, guided household discussions | 90-120 FGD participants, 7 households |
| [51] | Treatment of Childhood Diarrhoea in Nigeria: Need for | Nigeria | 2000 | Two sex stratified focus group discussions, 2 mixed | 32-48 participants |
|  | Adaptation of Health Policy and Programmes to Cultural Norms |  |  | sex focus group discussions |  |
| [52] | Childhood diarrhoea in a district in northeast Thailand: Incidence and treatment choices | Thailand | 2006 | Focus group discussions, in-depth interviews, surveys | 3 focus groups, 121 IDIs |
| [53] | Elder authority and the situational diagnosis of diarrheal disease as normal infant development in northeast Thailand | Thailand | 2009 | Semi-structured, open-ended interviews | 64 interviews |
| [54] | Predictors of diarrheal mortality and patterns of caregiver health seeking behavior in Karachi, Pakistan | Pakistan | 2016 | Focus group discussions | 29 participants (29/29 female) |
| [55] | Perspectives on child diarrhoea management and health service use among ethnic minority | Vietnam | 2011 | Semi-structured in-depth interviews, observations, focus group discussions | 43 interviews, 3 FGDs (0/24 female) |

|  |  |  |  |  |  |
| --- | --- | --- | --- | --- | --- |
| [56] | Environmental determinants of E. coli, link with the diarrheal diseases, and indication of vulnerability criteria in tropical West Africa (Kapore, Burkina Faso) | Burkina Faso | 2021 | Semi-structured interviews | 31 interviews |
| [57] | Navigating multiple options and social relationships in plural health systems: A qualitative study exploring healthcare seeking for sick children in Sierra Leone | Sierra Leone | 2014 | In-depth interviews, focus group discussions | 36 focus group participants (24/36 female) + 1 follow-up, 68 IDIs (68/68 female) + 2 follow-up |
| [58] | Influences on healthcare-seeking during final illnesses of infants in under-resourced South | South Africa | 2011 | Semi-structured interviews, key-informant in-depth interviews | 39 interviews (39/39 female), 19 key informant IDIs |

|  |  |  |  |  |  |
| --- | --- | --- | --- | --- | --- |
| [59] | Access to integrated community case management of childhood illnesses services in rural Ethiopia: A qualitative study of the perspectives and experiences of caregivers | Ethiopia | 2016 | In-depth interviews, focus group discussions | 78 IDIs, 132 FGD participants (132/132 female) |
| [60] | Operationalising integrated community case management of childhood illnesses by community health workers in rural Haryana | India | 2018 | In-depth interviews, focus group discussions | 37 focus groups participants (37/37 female), 43 IDIs (43/43 female) |
| [61] | Circumstances of post-neonatal deaths in Ceara, Northeast Brazil: Mothers' health | Brazil | 2000 | Semi-structured interviews | 127 interviews (127/127 female) |

|  |  |  |  |  |  |
| --- | --- | --- | --- | --- | --- |
| [62] | Treatment-seeking and recovery among young undernourished children post-hospital discharge in Bangladesh: A qualitative study | Bangladesh | 2022 | Repeat in-depth interviews, key informant interviews, focus group discussions | 73 repeat IDIs<br>15 Key informant interviews<br>18 FGD participants |
| [63] | Knowledge of, attitudes toward, and preventive practices relating to cholera and oral cholera vaccine among urban high-risk groups: Findings of a cross-sectional study in Dhaka, Bangladesh | Bangladesh | 2013 | In-depth interviews | 30 interviews (15/30 female) |
| [64] | “This diarrhoea is not a disease ...” local illness concepts and | Yemen | 2014 | Focus group discussions | 31 FGD participants (31/31 female) |

|  |  |  |  |  |  |
| --- | --- | --- | --- | --- | --- |
| [65] | A qualitative study of<br>community perceptions<br>about childhood<br>diarrhea and its<br>management in Assosa<br>District, West Ethiopia | Ethiopia | 2014 | In-depth interviews,<br>focus group<br>discussions | 72 participants<br>(60/72 female) |
| [66] | Comparing reports of<br>health-seeking<br>behavior from the<br>integrated illness<br>history and a standard<br>child morbidity survey | Egypt | 2008 | Interviews that<br>combined techniques<br>from in-depth<br>interviewing and<br>standard survey<br>interviewing | 137 participants |

<sup>1</sup>Information on sex of respondents provided only when available in text.

### Thematic Analysis

For clarity, we describe key insights from the thematic analysis linearly in the sections below. However, in reality care-seeking decisions are multifaceted and deeply contextual. Across the included studies, care-seeking behaviors were influenced by sociocultural practices and norms, perceived severity of the diarrheal illness, ease of access to different providers and facilities, and the perceived quality of those providers and facilities (Fig 2). The presence of themes, as well as their influence on care-seeking decisions, varies across studies. To explore how these themes affect care-seeking decisions overall and within individual studies, please refer to Table S2. A complete summary of all the themes and sub-themes that we identified are summarized in Table 2, along with the studies that described them.

**Fig 2.**
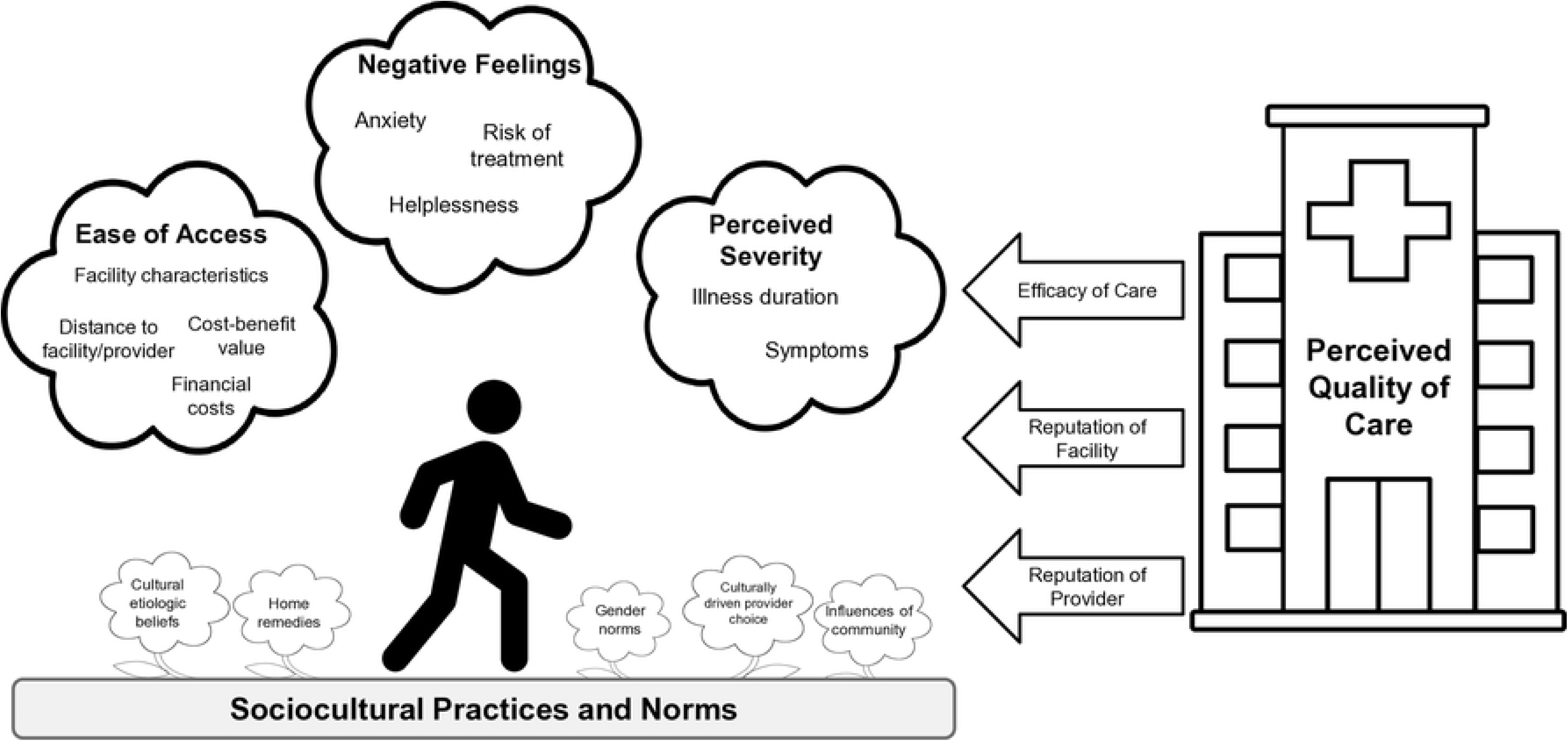
Visualization of care-seeking decisions. Factors that influence individual decisions to seek care across geographic settings. Diagram structure was adapted from Tefera and Yu [67].

**Table 2.**
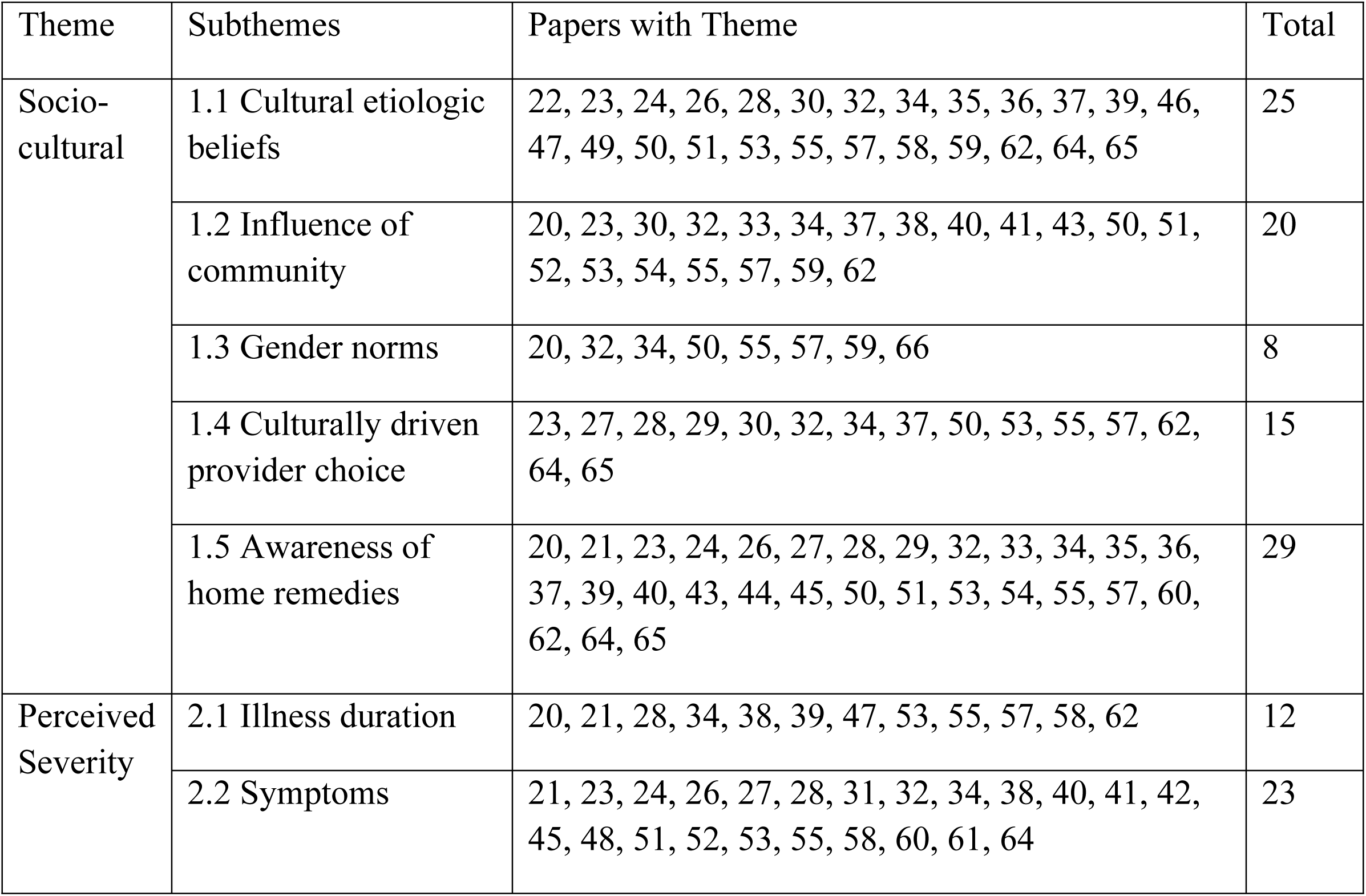
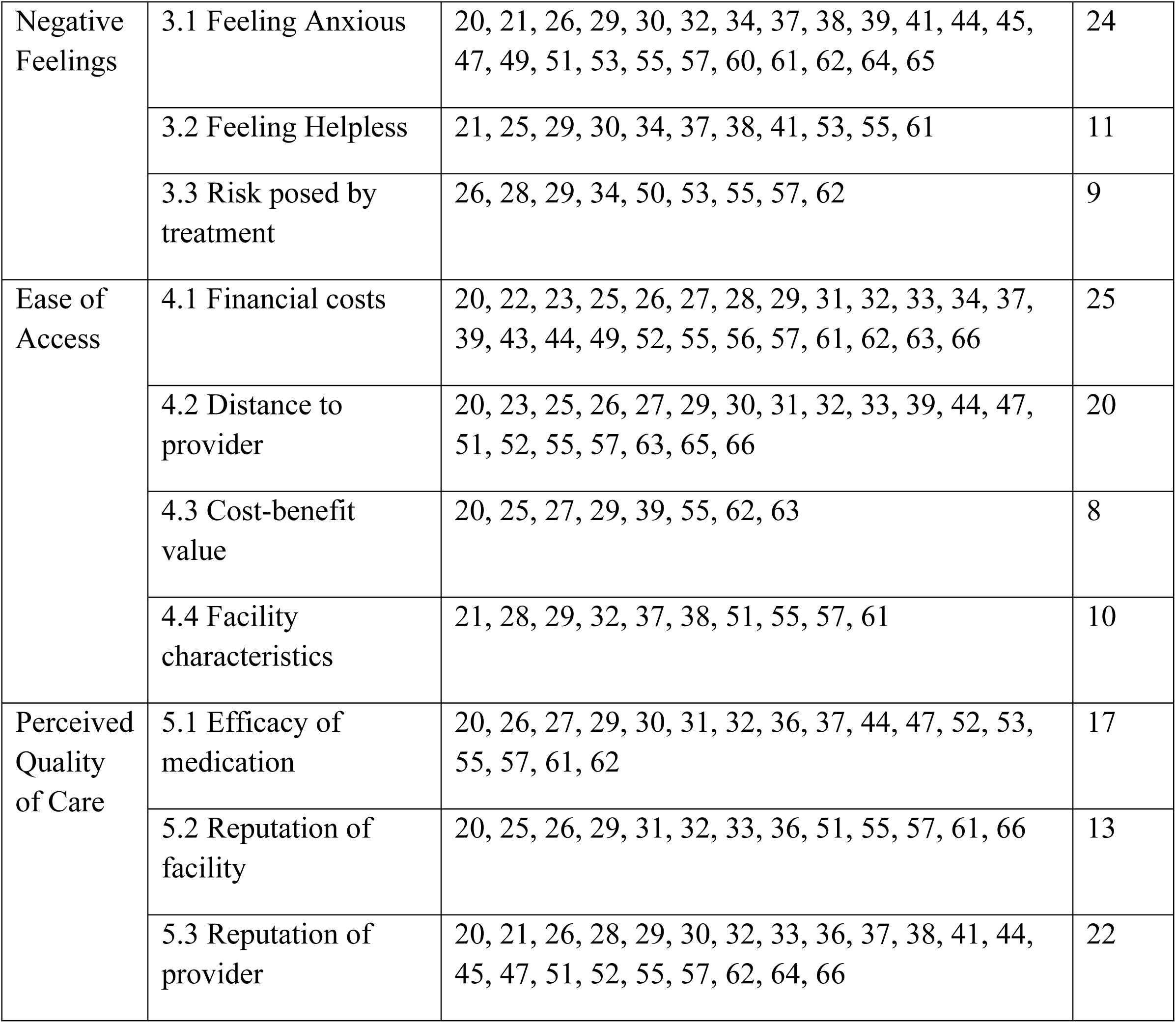
Frequency of Themes.

| Theme | Subthemes | Papers with Theme | Total |
| --- | --- | --- | --- |
| Socio-cultural | 1.1 Cultural etiologic beliefs | 22, 23, 24, 26, 28, 30, 32, 34, 35, 36, 37, 39, 46, 47, 49, 50, 51, 53, 55, 57, 58, 59, 62, 64, 65 | 25 |
|  | 1.2 Influence of community | 20, 23, 30, 32, 33, 34, 37, 38, 40, 41, 43, 50, 51, 52, 53, 54, 55, 57, 59, 62 | 20 |
|  | 1.3 Gender norms | 20, 32, 34, 50, 55, 57, 59, 66 | 8 |
|  | 1.4 Culturally driven provider choice | 23, 27, 28, 29, 30, 32, 34, 37, 50, 53, 55, 57, 62, 64, 65 | 15 |
|  | 1.5 Awareness of home remedies | 20, 21, 23, 24, 26, 27, 28, 29, 32, 33, 34, 35, 36, 37, 39, 40, 43, 44, 45, 50, 51, 53, 54, 55, 57, 60, 62, 64, 65 | 29 |
| Perceived Severity | 2.1 Illness duration | 20, 21, 28, 34, 38, 39, 47, 53, 55, 57, 58, 62 | 12 |
|  | 2.2 Symptoms | 21, 23, 24, 26, 27, 28, 31, 32, 34, 38, 40, 41, 42, 45, 48, 51, 52, 53, 55, 58, 60, 61, 64 | 23 |
| Negative Feelings | 3.1 Feeling Anxious | 20, 21, 26, 29, 30, 32, 34, 37, 38, 39, 41, 44, 45, 47, 49, 51, 53, 55, 57, 60, 61, 62, 64, 65 | 24 |
|  | 3.2 Feeling Helpless | 21, 25, 29, 30, 34, 37, 38, 41, 53, 55, 61 | 11 |
|  | 3.3 Risk posed by treatment | 26, 28, 29, 34, 50, 53, 55, 57, 62 | 9 |
| Ease of Access | 4.1 Financial costs | 20, 22, 23, 25, 26, 27, 28, 29, 31, 32, 33, 34, 37, 39, 43, 44, 49, 52, 55, 56, 57, 61, 62, 63, 66 | 25 |
|  | 4.2 Distance to provider | 20, 23, 25, 26, 27, 29, 30, 31, 32, 33, 39, 44, 47, 51, 52, 55, 57, 63, 65, 66 | 20 |
|  | 4.3 Cost-benefit value | 20, 25, 27, 29, 39, 55, 62, 63 | 8 |
|  | 4.4 Facility characteristics | 21, 28, 29, 32, 37, 38, 51, 55, 57, 61 | 10 |
| Perceived Quality of Care | 5.1 Efficacy of medication | 20, 26, 27, 29, 30, 31, 32, 36, 37, 44, 47, 52, 53, 55, 57, 61, 62 | 17 |
|  | 5.2 Reputation of facility | 20, 25, 26, 29, 31, 32, 33, 36, 51, 55, 57, 61, 66 | 13 |
|  | 5.3 Reputation of provider | 20, 21, 26, 28, 29, 30, 32, 33, 36, 37, 38, 41, 44, 45, 47, 51, 52, 55, 57, 62, 64, 66 | 22 |

### Theme 1: Sociocultural practices and norms guide care-seeking behavior

We found that cultural factors influenced perceptions of diarrheal illness and its appropriate treatment. While included studies represented many diverse settings, several common patterns of care-seeking emerged. Respondents widely reported integrating both biomedical and traditional approaches, blending elements of each rather than strictly following either. These beliefs influence how symptoms are understood, who is respected as the authority of the illness, and thus how respondents make care-seeking decisions. This syncretic system reinforces the importance of culturally competent care and community-based participatory research.

#### 1.1 Cultural etiologic beliefs

We found that cultural beliefs about the cause of diarrhea often influenced care-seeking behavior. Study participants in ten studies made a distinction between diarrheal episodes that were recognized as diarrheal illness and diarrhea that was not associated with any illness. Caregivers commonly viewed diarrhea as a normal part of childhood development, not as a symptom of disease, but as a normal part of childhood development. As explained by one mother in urban Yemen, “This diarrhoea is not a disease; it never responds to doctors’ advice” [64]. These episodes were seen as benign and self-limiting, not requiring treatment while other forms of diarrhea were interpreted as signs of disease that necessitated treatment outside the home [22,23,32,37,39,46,51,53,58,64].

When diarrheal episodes were seen as pathogenic, perceived causes shaped both how severe it was perceived and treatment choices. Caregivers believed some illnesses were “traditional” and untreatable with Western medicine. These illnesses, though recognized as pathogenic, were considered uniquely specific to the community and distinct from modern disorders, requiring traditional rather than medical treatments. Traditional illnesses were often viewed as serious and required traditional rather than medical treatments [22,32,34–37,39,46,47,53,55,57,64]. Similarly, perceived supernatural causes of diarrhea could deter families from seeking care at a hospital or clinic, leading them to consult traditional or spiritual healers instead. As these cases were typically regarded as serious, treatment was usually sought promptly. Respondents’ belief in witchcraft, punishment from the gods, evil eyes, malicious spirits, and undefined supernatural entities were not limited to any specific region [24,26,28,30,35,37,39,46,47,49,50,58,59,62]. In contrast, when diarrhea was attributed to weather or diet, the illness was often viewed as mild and self-limiting, thus not requiring care from hospitals or clinics [23,28,30,37,39,46,50,51,55,58,59,62,64,65]. As a mother in South Africa explained, “I did not think it was serious. I thought it was due to cold weather and it will subside” [58].

#### 1.2 Influence of community

Community members played a wide variety of roles in care-seeking including giving advice, herbs for treatment, financial or logistical support, or even permission if a father or husband was not available. Elders in rural communities often discouraged the use of modern medicine in favor of traditional treatments in sub-Saharan Africa and Southeast Asia [20,23,30,32–34,37,38,40,43,50–55,57,59,62]. As one parent stated, “Everything must be discussed with the grandparents […]. After discussing, if all agree, we let the child go (to the CHS)” [55].

#### 1.3 Gender norms

We also found that gender roles in decision making shaped care-seeking. In five studies, fathers held financial control and authority within families, especially in rural sub-Saharan Africa. Women participating in the included studies often recounted having limited autonomy to seek care for their children without first obtaining their husband’s approval, which could delay treatment. If the male authority figure was unavailable or unwilling to provide money for care, women turned to traditional medicine or informal vendors as cheaper alternatives [20,32,34,50,55,57]. As a father in Nigeria explained, “Money can actually delay the mother from seeking care; as a mother when you don’t have money, you would have to wait for the father…” [32]. In one of the rare studies that did not limit the inclusion to mothers, fathers in a rural area of Ethiopia were deterred from seeking care at health posts, which respondents described as women-only spaces. One father explained, “I do not go in the health post as it is only for women and children.” Fathers also reported not knowing what treatments or levels of care were available at these posts [59]. A study conducted in Egypt during 1996-1997 found that a child’s sex influenced care-seeking behavior. As one mother explained, “Here the attention is more focused on the boy than the girl. For example, if the boy gets sick, we take him to the doctor, but if it is the girl… God supports her, and she gets well on her own.” [66].

#### 1.4 Culturally driven provider choice

Cultural norms often shaped care-seeking practices, not always through specific beliefs but because it was simply what one does: what is familiar and expected [23,27–30,32,34,37,50,53,55,57,62,64,65]. The choices often reflected a sense of embeddedness in cultural routines and expectations, where traditional care was not necessarily weighed against biomedical options, but accepted as a part of the natural course of action. As one mother in rural South Africa explained, both options exist within that familiar world, “There is help from a traditional healer and there is help from the hospital so you don’t know where your child is going to get help. And there are things that sangoma [traditional healer] can do and the hospital cannot do and there are things that the hospital can do that a sangoma cannot do” [30].

#### 1.5 Awareness of home remedies

Home remedies were a culturally ingrained part of care-seeking behavior in many communities. Reliance on home remedies could act as both a solution and a barrier [20,21,23,24,26–29,32–37,39,40,43–45,50,51,53–55,57,60,62,64,65]; they may provide immediate relief or comfort for mild or moderate diarrhea, but study authors noted it could also delay medical care [20,27,37,44,54,62].

### Theme 2: Diarrhea perceived to be severe is treated outside the home

We found that care-seeking for diarrheal disease can be viewed on a continuum that varies with diarrhea severity. Respondents often reported higher rates of care-seeking for diarrheal illnesses they perceived to be more serious, while illness episodes viewed as minor were primarily treated with home management or traditional remedies. Participants also utilized illness duration and symptoms as tools to assess the severity of the illness and a means to determine what, if any, type of care-seeking was necessary.

#### 2.1 Illness duration

Participants in the reviewed studies often described escalating care-seeking depending on the length of symptoms, moving from home management to outside care or traditional healers to health facilities, with the timeframes for these transitions varying between studies from days to years [20,21,28,34,38,39,47,53,55,57,58,62]. For example, among caregivers in urban Canada, when their child’s illness lasted longer than they expected, they escalated care-seeking to the pediatric emergency room [38].

#### 2.2 Symptoms

Study participants viewed the presence or absence of certain symptoms as evidence of severe disease. Signs of dehydration [21,24,26,32,42,55,60], diarrhea in combination with fever or vomiting [38,51–53,60,61], decreased or loss of appetite [42,51,61], and bloody or watery diarrhea [23,24,26,42,45,51,52,64] were all noted as signs that participants should seek care outside the home for their child. Specifically, caregivers that identified signs of dehydration [21,24,26,32,41,42,55,60] or symptoms consistent with cholera [23,24,26,42,45,51–53,64] largely reported seeking care outside the home. Respondents frequently did not consider diarrhea on its own severe and thus it was not treated outside the home [27,28,31,34,40,48,51,58]. Canadian respondents reported conflicting influences of observed ‘suffering’ during diarrheal illness on care-seeking, varying by age. For caregivers of children in a mixed urban rural setting, the perception their child was suffering led them to the emergency room [38] while for adults in a rural community of indigenous people in Canada, acute gastrointestinal illness was something the community believed a person had to “suffer through” [40].

### Theme 3: Negative feelings drive the type of care sought

Study participants who used modern health care providers often expressed negative feelings, including anxiety, helplessness, and fear. These negative feelings collectively influenced respondents’ healthcare decisions, reflecting how emotional state drives the nature and urgency of care sought. This sense of urgency often led to care-seeking in hospitals or clinics. For others, fear over the treatment itself might result in avoidance of hospitals or clinics.

#### 3.1 Feeling anxious

Worry was a common reason caregivers sought care outside the home. Concern over a patient’s vulnerability, due to their age or pre-existing conditions, often prompted seeking more intensive care, such as moving from home remedies to clinical treatment or from primary care to emergency care. Patient vulnerability frequently led to an increased sense of urgency [21,30,38,41,45,53,55], as one caregiver in Canada explained “I didn’t want it to go on to a point where it was going to cause more problems for her so I wanted to get her in as soon as possible” (Interview 4) [38]. In many cases, caregivers described seeking more advanced levels of care when the illness did not improve or worsened, as continued symptoms deepened their concern and prompted action [20,21,26,29,32,34,37,39,44,47,49,51,57,60–62,64,65]. Feeling distress over the uncertainty of what was wrong was also an impetus to seeking care [21,38].

#### 3.2 Feeling helpless

Feelings of helplessness in managing diarrheal illness prompted caregivers to seek care and reassurance from healthcare providers [21,25,30,34,37,38,41,53,55,61]. This was especially true for first time parents [34,38,53,55], including a caregiver in Canada who stated “I tried to [treat the illness] this time…I really try my best to take care of what I can here, and then if things get to a point that I’m a little worried, then we normally go in to see a doctor” [38]. Caregivers also described feeling helpless if they felt they had limited options for care outside of the home [29].

#### 3.3 Risk posed by treatment

Participants, especially caregivers of sick children, sometimes viewed Western treatments intended to help manage diarrheal illness as harmful. Participants reporting these concerns came from rural areas [26,28,34,50,53,55,57] or had arrived in an urban area from either slums or rural areas [62]. Participants reported fear that the Western medicine and pharmaceuticals offered might be harmful, and possibly even cause death. This led to decreased use of health facilities and drug peddlers whom caregivers did not trust [26,28,45,50,55,57,62,64]. One mother stated that she finds “herbal medicines more ‘compatible’ for a body of a child since they are not harmful as drugs can be" [55]. In rural Uganda, respondents were concerned that hospitals could expose them to other pathogens [29].

### Theme 4: Ease of access shapes care-seeking practices

Structural and logistical factors played a critical role in shaping participants’ ability to obtain healthcare. Financial burden, including both direct service costs and indirect expenses, were key deterrents. Many participants also reported barriers, such as distance and operating hours, that diminished their ability to seek care at desired facilities. These barriers often create opportunity costs, as overcoming them requires time, energy and financial resources that could be allocated to other essential needs. This dynamic may reflect systemic inequities that compound barriers to care, making the decision to seek treatment even more challenging.

#### 4.1 Financial costs

Costs were frequently mentioned as a reason to avoid or delay care at a hospital or clinic. This often led participants to rely on traditional healers, traditional medicines, or informal drug sellers because they were less expensive [20,22,23,25–27,29,31–34,37,43,44,49,55–57,61,63,66]. As a respondent in rural Bangladesh explained, “It is not easy for us to seek medical treatment now, it is easy to seek treatment when you have the money and can afford to access the medical facilities available’’ [33]. Similarly, a participant in rural Uganda explained that “If I have not paid the hospital, I try by all means to get herbs from forests or places around my home. I take the herbs, and if they fail to heal me, I wait for death (male)” [29]. In addition, one study reported that pharmacists or informal service providers that offer payment plans were preferred when participants could not pay all hospital fees up front [32].

Specific medical costs associated with not seeking care included the cost of medications, consultations, admission fees, and/or laboratory tests. Studies mentioning these costs were all in rural areas [20,25,27,32–34,44,55,61,63], although some of these locations noted having free services or fee exemption programs in place for children under five [20,25,32,44,55,61]. Only urban caregivers in a mixed study population in Kenya mentioned free or low-cost care being available, and it was those caregivers who stated they frequented health facilities when their children were sick [26]. Caregivers in Thailand reported occasionally using a health center because it was free, but most often used private services, despite the increased cost, because they found that the services were better [52]. Eleven studies, all in rural areas, reported transportation costs associated with care-seeking were a barrier to seeking care at formal care providers [20,25,29,32–34,37,39,55,57,62,63].

#### 4.2 Distance to provider

Distance to a health facility was frequently reported as a barrier to seeking care, especially in studies in rural areas. Distance often led to decreased ability to get to clinics, hospitals, and other medical providers and so respondents relied instead on traditional healers, drug sellers, and home remedies [20,23,25–27,29–33,39,44,47,52,55,57,65,66]. However, what constituted distance varied widely and was not consistently reported by study authors. In South Africa, one caregiver described traveling 60 kilometers to obtain medication for their child as easy [30] while another recounted the half to full day walk as a huge barrier to seeking care in rural Vietnam [55]. As explained by one mother in Nepal, “I believe in herbs but not in traditional healers. However, my mother took my baby to dhami-jhankri, a traditional healer and she was cured. If hospital is far, we should sometimes visit traditional healers for the sake of child” (IDI-mother 2) [23]. Many respondents mentioned needing to walk to the health care provider. This was even more challenging in flood-prone areas, where travel by foot or vehicle was frequently impossible or difficult to navigate [25,39,44,51,58]. Many caregivers described selecting whoever was closest to home, as explained by one focus group participant, ‘if the health centre was close by, we would go there for medication, but we always find troubles getting to Bwindi Hospital, so we have to take herbs’ [29]. Another mentioned a ‘fear of walking long distances’ and that it ‘always takes a lot of time to reach the hospital’ [29]. In a study that interviewed respondents from both urban and rural areas, some caregivers sought care from health facilities and chemists because they were closer to their homes. However, rural community members still cited distance as a barrier [26].

#### 4.3 Optimizing cost-benefit value

Some studies found that the direct and indirect financial burden of care-seeking necessitated forgoing other important expenditures such as food [27,55,63] and clothing [29,62]. Seeking care outside the home might also require taking time off from work which again reinforced the financial burden of care-seeking [20,39]. Respondents, particularly female caregivers, cited competing priorities–such as work obligations, childcare, and household needs–that prevented them from seeking care outside the home [20,25,39,55,62]. Participants weighed the costs of transportation and services against the potential benefits of care, which were not always certain. This was illustrated by a mother in rural Kenya, who stated “sometimes, even if you find money and go [to the health facility] you feel like you are wasting money because it [treatment] doesn’t always work” [25].

#### 4.4 Facility characteristics

Limited facility hours resulted in delayed care, especially when illnesses were identified at night when hospitals might not be open. In these situations, caregivers described relying on traditional medicine and healers, drug peddlers, family, and home care [28,32,51,57,61]. Caregivers described prolonged wait times as a reason to avoid certain facility types [21,28,29,32,37,51,57]. As Brubacher et al. described, “We always get to the hospital around 8am, but we spend the whole day not being attended to, and sometimes they close up when they have not attended to us. This doesn’t make us happy at all. In most cases, when that happens, I never go back to the hospital, but go back home and look for my herbs (female)” [29]. Only among ethnic minority caregivers in rural Vietnam did language barriers impact care, as facilities did not provide translators, requiring those seeking to bring someone with them or rely on patients at the facility to act as translators [55].

### Theme 5: The perceived quality of care influences care-seeking

We also found that the decision to seek care outside the home was influenced by participants’ perception about the efficacy of treatment and the reputation of the facility or provider. While many respondents’ explanations of how sociocultural factors influenced the types of care sought, respondents in many settings also reported viewing modern pharmaceuticals as essential for treating diarrhea, and often sought out facilities with reputations of being well stocked. Quality issues were mentioned in 27 unique studies, 25 of which also mentioned sociocultural factors.

The actual capacity of a health care site to deliver care was important and decisions were strongly shaped by past experiences that built trust and confidence in providers’ competence. This illustrated how those seeking care must weigh the tangible benefits of medical treatments and their personal relationships with those providing it.

#### 5.1 Efficacy of medication sought

Respondents frequently noted the efficacy of modern pharmaceuticals as a reason for seeking care outside the home, especially in Sub-Saharan Africa and Southeast Asia [20,26,27,29–32,36,37,44,47,52,55,57,61,62]. For example, a caregiver in rural Uganda stated, “We always take the drugs and get cured…we always take the first option of going to a hospital” (female) [29]. Some believed the medications would stop the diarrhea [26,29,36,37,52,55], while others mentioned it as a cure [29,37]. As explained by a caregiver in Bangui, Central African Republic, “When you give the child medicine…this will stop the diarrhea. Diarrhea, isn’t that an illness? If you don’t give the child medicine, she won’t get better!” [37]. This positive view of medications resulted in the selection of private facilities farther from home for study participants in several studies [20,36,52,53].

#### 5.2 Reputation of facility

Study respondents reported that knowing a facility was adequately equipped with both doctors and medications positively impacted their decision to seek care there [20,25,26,31–33,36,51,55,57,61,66]. When medications were not available at health facilities, respondents reported visiting informal providers or delaying care-seeking [20,31,33,51,55,57,61]. Participants in several studies noted that availability of medication was most frequently a problem at government and public health facilities [20,25,36,51]. We identified only one study among an indigenous community in rural Uganda that stated the cleanliness of the health facility and its improper maintenance was a barrier to seeking care there [29].

#### 5.3 Reputation of provider

We found that the reputation of a provider or facility for providing effective treatment engendered trust and confidence in seeking care there [20,29,30,32,36,38,41,47,52,57,62]. Reasons for trusting providers included a history with the facility [30,36,57], successful treatment of past diarrheal illness [38,47], or a high quality service/interpersonal relationship with the provider [32,44,57]. Others stated a more general feeling of confidence in the providers’ competence [21,29,32,38,40,57]. Confidence that the provider could heal a child was especially important, as explained by a respondent in Canada, “…you know, it’s [the hospital emergency department] a magical place. Next time I’m bringing her after one day because right after we go, it always works out the same, [the illness] stops” [38]. Cunnama and Honda described trust in hospital providers as a newer phenomenon, with one participant stating that “…it’s a good hospital…nowadays it’s not like before when our mothers used to go there. Like now, everything is done well there. It’s the right hospital.” [30]. Five studies, in both rural and urban areas, mentioned the desire to get a diagnosis as a reason to seek care at modern health facilities [20,21,26,32,55,57]. Previous experiences of poor treatment, such as failure by the provider to successfully treat a past illness episode, providers responding negatively to patient accounts of home treatments administered before arrival or refusing to listen to patients’ understanding of the illness, also prevented caregivers from visiting specific health providers [20,28,33,37,45,51,55,57,64,66].

## Discussion

This review identified several barriers and facilitators to seeking care outside the home for diarrheal illness. We found that socio-cultural beliefs about illness and caregiver response, the perceived severity of the illness, emotional reactions to a child being sick, accessibility of care, and the expected quality of care all impacted the decision to seek care outside of the home across diverse geographic locations. In addition, while reasons for or against seeking care outside the home were presented here as discrete factors, respondents often reported multiple influences—some reinforcing each other and others in conflict—shaping their decisions. For example, an elder in the home advocating for traditional methods influenced decision-making, even in the presence of concerns about disease severity or etiologic cause [50,53,59]. Although respondents in one study trusted the quality of care available at a hospital, financial hardships remained a barrier to utilization [37]. Proximity to government facilities was not a facilitator of care-seeking when medications and equipment were unavailable, facilities were closed on weekends, or providers were reported absent [51]. Even positive views on the efficacy of medication are overlooked when home remedies and traditional healers are familiar and proximate [57]. Ultimately, what an individual would have liked to do did not always align with what they could do.

In addition, we found that general diarrhea—without signs of dehydration or blood in the stool—was often viewed as a routine part of child development rather than an illness requiring medical attention. This was frequently cited as a reason for not seeking care outside the home and may partially explain why we found no significant differences in hospital care-seeking for children under five compared to older individuals in our previous review of population-based survey data [15]. While many examples related to mild episodes that may not require care outside of the home [22,23,32,35,37,51,55], some involved moderate-to-severe dehydration or were described as illnesses that lead to extended hospital stays or death [24,28,34,50,53,57,64]. Considering these differences in understanding diarrhea, using local terminology for diarrheal episodes and including descriptions of the event (i.e. three loose stools) could improve accuracy of studies investigating prevalence and care behavior for diarrheal illness.

We also found that traditional healers were more commonly reported as a source of care in the qualitative studies than in our previous review of quantitative studies [15]. This difference may reflect the fact that non-medical providers are included less frequently in survey questionnaires [15] and/or could be a result of how the questions themselves are phrased. For example, participants may be less likely to report use of a traditional healer if only one source of care can be selected. Additionally, different perceptions of illness may influence responses to survey questions, as respondents in qualitative studies differentiated between illnesses requiring biomedical treatment and those best addressed by traditional or spiritual healers. Not explicitly asking about behaviors for a broad range of diarrheal symptoms may lead to incomplete utilization histories, particularly for traditional and spiritual healers.

Our findings closely align with several established theoretical models identified by a review of healthcare utilization models, particularly those by Andersen [68,69] and Kufafka et al.’s [70] frameworks. Both models emphasize the importance of health beliefs, access to healthcare, social support, and perception of illness. Cultural beliefs and severity of illness influence how individuals view their health and the need for care, much like Anderson’s model suggests. Fear, as identified in our review, ties into the emotional and psychological barriers highlighted by Kufafka et al., which stress the role of emotional responses, symptom interpretation, and self-efficacy in the decision-making process. Together, these models and our findings point to a range of internal, external, and community-level factors that collectively shape whether and how individuals seek care [71].

While these findings provide context for understanding care-seeking behavior, several limitations should be acknowledged. First, this is a synthesis of data which previously underwent synthesis. It is possible that loss or alteration of information occurred in either or both stages. Second, there was great variety in study designs, care provider options described and illness definitions, limiting direct comparison between studies. While no restrictions were set on study population or geographical location, the majority of studies were conducted in caregivers of children in Sub-Saharan Africa and Southeast Asia, which may limit the generalizability of findings to other settings. Despite the rigor of our methods, it is possible the search strategy and/or screening process missed pertinent articles. Studies missed may have produced stronger or contrary information than is presented here. Finally, all authors of this study are from high-income countries, whereas most studies were conducted in low-income countries or low-resource settings. While we have aimed to center participant perspectives, our analyses and conclusions may still be influenced by implicit biases shaped by our backgrounds.

Our findings highlight key methodological gaps that could be addressed in future studies. While there was broad consensus that severe illness prompts urgent care outside the home, it was not always clear how respondents defined severe illness. Including clear and location-appropriate case definitions and symptom descriptions will be important for future work. Consistent with our previous review [15], community health workers rarely were mentioned as a source of care. It remains unclear if this is evidence of nonuse or a result of how “healthcare” is defined. As community health workers are trained community members themselves rather than medical professionals [72,73], they may not be thought of as official sources of care without direct prompting. This perspective is supported by Hodgins et al., who note that the term community health worker encompasses a diverse cadre of roles and responsibilities, which complicates consistent recognition of their role as formal healthcare providers [74]. Finally, this review highlights the relative scarcity of qualitative studies compared to quantitative research on care-seeking for diarrheal illness. Given the wide variation in care-seeking across settings [15], mixed-methods approaches that examine where, when, and why people seek care will be important for understanding local-level care-seeking.

The themes we identified as shaping care-seeking decisions across diverse settings offer important insights for designing both interventions and more responsive surveillance systems. Understanding how and when individuals seek care can help identify where case detection is likely to miss milder illness, where mistrust or limited access hinders reporting, and where community-based approaches may be needed to better estimate disease burden, despite their resource demands. In addition, expanding educational campaigns that engage with local understandings of disease etiology may support more timely and appropriate care-seeking. This review found that decisions were influenced more by perceived necessity than by cost alone, suggesting that interventions should focus on improving recognition of when and what type of care may be needed. For these efforts to succeed, individuals must also have trust in local healthcare providers. Depending on the context, this may involve ensuring providers have appropriate treatments, adequate staffing, or engaging community health workers and trusted local leaders in care delivery. Strengthening caregiver knowledge and confidence in a setting-specific manner may help reduce delays or avoidance in seeking care, ultimately contributing to reduced morbidity and mortality from diarrheal disease.

## Data Availability

Qualitative codebooks, study characteristics, and themes identified in each study are available within the manuscript and associated files.

## Supporting information captions

**S1 Data. Study Characteristics Dataset.** Excel sheet with study characteristics extracted from all studies that met inclusion criteria (tab 1). Tab 2 shows variable descriptions.

**Table S1. Thematic Codebook.** Coding framework developed and applied in Atlas.ti during the initial coding phase. Parent codes represent anticipated broad thematic categories, while child code captures more specific sub-concepts.

**Table S2. Theme Influence on Care-Seeking by Study.** Theme presence and influence on care-seeking behavior across studies, based on quotes coded in Atlas.ti.

